# Statistical Analysis of Pre-War Primary Healthcare Costs in Ukraine: Variations by Location and Ownership and Implications for Financing Reform

**DOI:** 10.64898/2026.07.15.26358144

**Authors:** Aqeel T Mohamed, Kaija Kasekamp, Olga Demeshko, Triin Habicht, Adrianna Murphy, Zia Sadique, Jarno Habicht

## Abstract

Strong primary healthcare (PHC) is associated with lower costs and better population health outcomes when supported by appropriate financing. Costing analysis enables evidenced-based decisions for estimating budgets for PHC and defining provider payments. In 2021, a project supported by the World Health Organization was launched in Ukraine to collect cost data from 100 PHC providers. The objective was to assess costs for delivering services within the state-funded benefits package, with the aim of informing tariff-setting, and assessing budget need.

This study used statistical analysis on the collected cost data. We applied multivariable linear regression (MLR) to assess variation in cost-per-person across locality (rural vs. urban) and ownership type (public vs. private) of the providers, after adjusting for confounders.

The mean (standard deviation) cost-per-person across the sample providers was 45.46 (18.46) USD. MLR analysis showed that rural providers had a higher cost-per-person of 6.70 USD (95% CI: 1.54, 11.85) compared to urban providers, after adjusting for confounding (p=0.011). We also found strong evidence that private providers had a lower cost-per-person of 36.15 USD (95% CI: −41.82,-30.48) compared to public providers, after adjusting for confounding (p<0.001).

Although our findings do not capture the impact of Russia’s hostile invasion of Ukraine, they still provide valuable insights for policy discussions within Ukraine and for other nations examining PHC financing reforms. Our findings align with international evidence suggesting that rural providers incur higher costs, supporting the need to adjust capitation payments for providers in these areas. Ownership type also affects costs, potentially reflecting differences in quality standards between public and private providers. These differences allow private providers to opportunistically reduce costs by limiting staff numbers and optimizing facility size to maximize profits. To ensure equitable access to high-quality PHC, uniform service delivery standards should be applied to all PHC providers, regardless of ownership type.

## Introduction

A strong primary healthcare (PHC) system provides essential, affordable and accessible care that addresses the health needs of entire communities, including health promotion, disease prevention, and health maintenance (1,2). Globally, PHC has been recognized as essential in achieving Universal Health Coverage and the Sustainable Development Goals (3,4). To fulfil its role, PHC requires appropriate financing that reduces out-of-pocket spending and supports equitable healthcare access (3). High out-of-pocket spending is a known barrier to accessing PHC and other types of care, and can lead to impoverishment, particularly among poorer households who typically have the greatest need for PHC (3,5,6).

In Ukraine, the Ministry of Health of Ukraine has prioritized strengthening PHC, as evidenced by the implementation of major reforms beginning in 2017 (7–9). The 2017 Law on Financial Guarantees for Healthcare Services mandates that all Ukrainian citizens are registered (signing a declaration) to PHC providers contracted by the National Health Service of Ukraine (NHSU). This ensures free access to services covered by a benefits package – the Program of Medical Guarantees (PMG). The NHSU acts as the sole national purchaser of publicly financed PHC services.

PHC providers eligible for NHSU contracts include three types: 59% public providers (local or central government owned), 13% private providers (limited liability companies) and 28% Fizychna osoba-pidpryemets (FOPs) in 2021 (10). FOPs are small private practices that employ mostly a single doctor, with some employing additional practitioners (8). The current Ukrainian PHC system is financed through an age-adjusted capitation-based system (i.e., providers being paid a fixed amount in advance per registered person to deliver the PHC services described in the PMG). There is also an adjustment of 1.2 for the capitation payments for providers serving mountainous areas (8).

Russia’s hostile invasion of Ukraine has led to reductions in public budgets and limited the public funding allocated to the NHSU (7,9,11). This has heightened the need to maximize ‘value for money’ by strategically paying for services and contracting providers. Despite these challenges, the Ukrainian government has made efforts to maintain public spending on PHC (11). Historically, Ukraine’s public budgets for PHC services were based on previous years’ spending rather than comprehensive needs assessments or bottom-up cost analyses, which hindered the ability to ensure adequate funding for service delivery.

To address this challenge and inform a revision of how PHC financing is determined, the World Health Organization (WHO), in partnership with the NHSU, conducted a PHC costing study from 2021 to 2023. The study collected and analysed data to inform PHC funding requirements and tariff setting (12). The study relied on pre-war cost data, but data collection occurred retrospectively after the invasion. The conflict impacted data availability as some providers (particularly those in close-conflict areas) were unable to share their data, limiting the sample (**Figure 1**). The analysis presented in this paper uses data from the PHC costing study to conduct a cost variation analysis, assessing the impact of locality and type of ownership (explanatory/independent variables) on cost-per-person (response/dependent variable) of PHC providers. The study aims to provide evidence on the difference in cost structure and level, providing valuable information for designing provider payments, as well as to define service delivery standards and develop contracting mechanisms for PHC services for Ukraine and also other countries seeking evidence on reforming PHC systems with the aim to promote equitable access to high quality healthcare.

**Figure 1:**
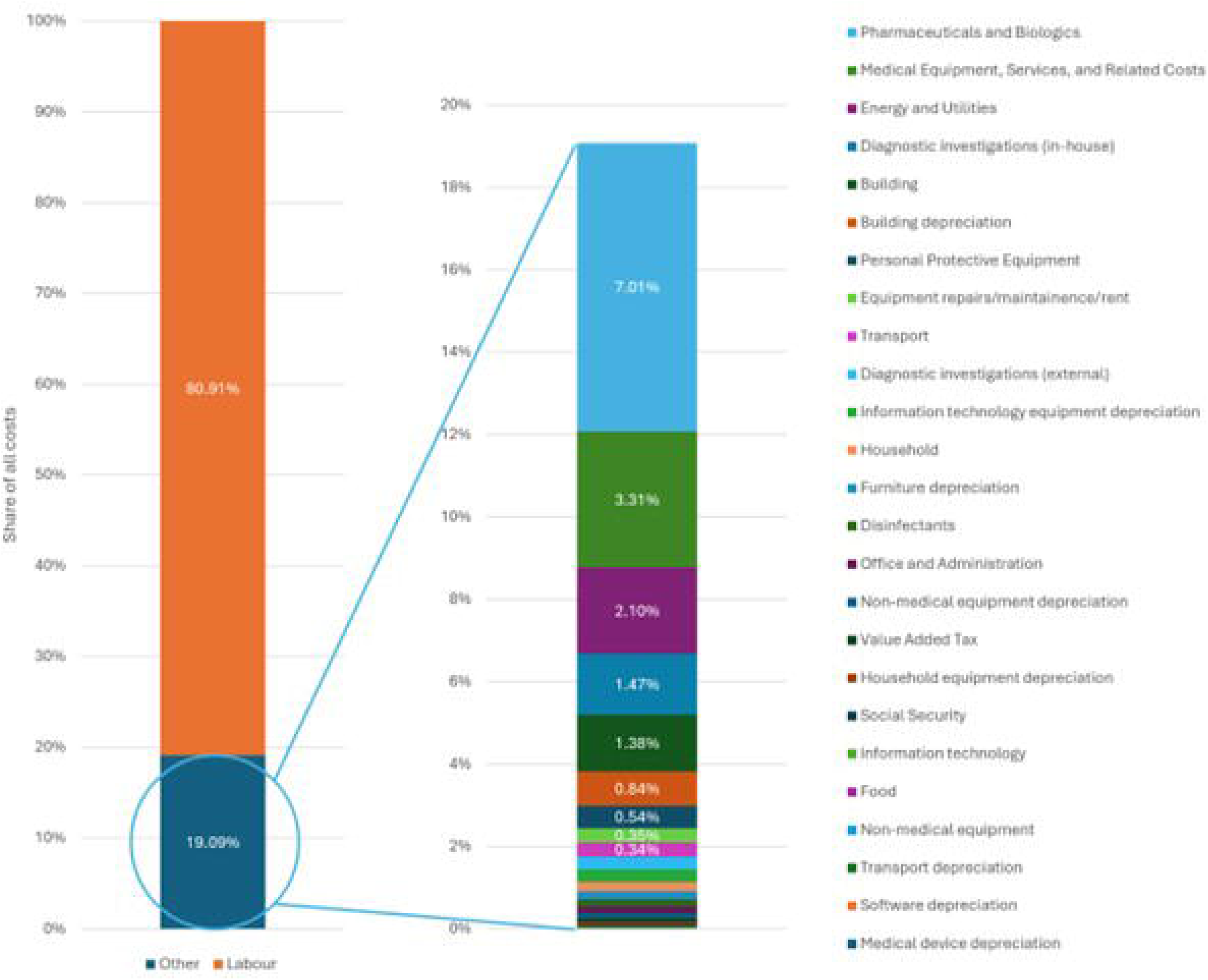
Geographical distribution across Ukraine of the 100 Primary healthcare providers involved in the costing study. Alt text: Map of Ukraine outlining the geographical distribution of Primary healthcare providers involved in the costing study. The map also shows the territories occupied by the Russian Federation before February 2022 along with territories that were occupied after February 2022 and liberated as of March 2023.

## Materials and Methods

We converted the PHC cost data from Ukrainian Hryvnia (UAH) to 2021 United States Dollars (USD) using the 2021 exchange rate provided by the Central Bank of Ukraine (1 USD = 27.29 UAH)(13). We conducted and reported all statistical analysis in USD.

This cost variation analysis used 2021 costing data from 100 Ukrainian PHC providers selected using convenience sampling. **Table S1** and **S2 (Supplementary Material)** outline the main methodological approach and inclusion criteria for the Ukrainian PHC costing study.

This analysis used cost-per-person as the primary response (dependent) variable. Using cost-per-person offered the advantage of standardized comparison of results between different PHC providers, regardless of the total number of registered persons they serve.

The equation used to calculate cost-per-person is outlined below:

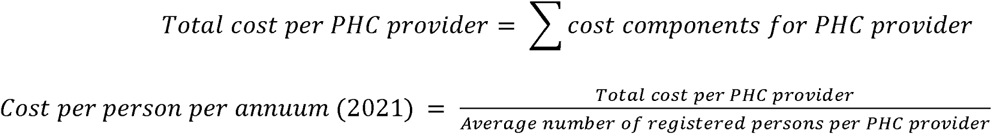

**Table S3 (Supplementary Material)** outlines the cost components that counted toward the total cost per PHC providers.

**Table 1** outlines the explanatory (independent) variables for this study.

**Table 1:**
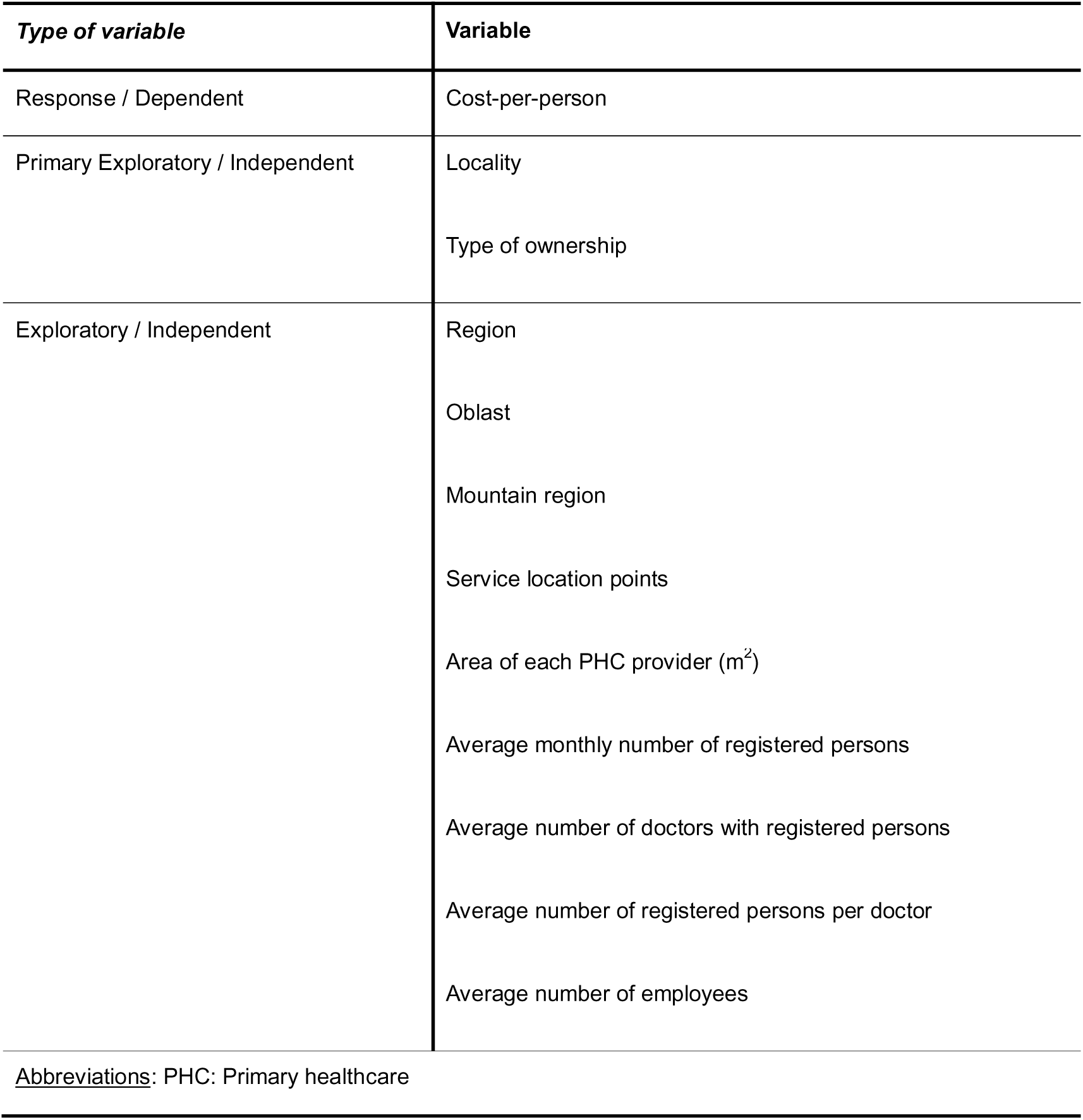
Definition and classification of study variables.

| *Type of variable* | *Variable* |
| --- | --- |
| Response / Dependent | Cost-per-person |
| Primary Exploratory / Independent | Locality<br>Type of ownership |
| Exploratory / Independent | Region<br>Oblast<br>Mountain region<br>Service location points<br>Area of each PHC provider (m<sup>2</sup>)<br>Average monthly number of registered persons<br>Average number of doctors with registered persons<br>Average number of registered persons per doctor<br>Average number of employees |
| <u>Abbreviations:</u> PHC: Primary healthcare |  |

We selected locality and ownership type as the primary explanatory variables, as both have evidence of influencing PHC costs (14–28). Preliminary descriptive analysis from the Ukrainian PHC costing study also found both variables to be associated with different levels of costs (8). Specifically, a significant body of evidence links rurality with incurring higher healthcare costs (14,20–23,25,26,29,30). However, the association between ownership type and healthcare costs remains less clear, with conflicting evidence in the literature (15–19,24,27,28).

The remaining independent variables were defined based on provider characteristics that may impact the costs. These included PHC provider region (defined for NHSU contracting as North, South, East, West or Central Ukraine), oblast, and whether a provider operated in high mountainous area. PHC providers in Ukraine also frequently had multiple service location points, which may influence costs. PHC costs may have also depended on the total number of persons registered, physicians serving the registered population, persons registered per physician, and the number of employees. We calculated the number of registered persons per provider, registered persons per doctor, doctors with registered persons, and the total number of employees by averaging their respective values across each PHC provider throughout 2021

We conducted this this statistical analysis to assess the impact of locality and ownership type (explanatory/independent variables) on the cost-per-person (response/dependent variable) of PHC providers.

Statistical analyses used Stata v18.0 and Microsoft Excel 2021 v2406.0. We set the alpha-level for this study at 0.05, following standard convention (31–34).

We used histograms, Q-Q plots and, the Shapiro-wilk test, to assess the normality of the cost-per-person variable. We chose the Shapiro-Wilk test for its perceived higher power and accuracy in evaluating normality (35,36).

We conducted preliminary descriptive statistical analysis to explore the baseline characteristics of the 100 PHC providers. We used the mean and standard deviation to represent central tendency and spread, respectively. Given the study’s large sample size (n=100), we applied the Central Limit Theorem (37–40), which validated the use of the mean and standard deviation (despite being parametric measures of central tendency and spread, respectively) (37,38,40). We also calculated numbers and corresponding percentages for each explanatory/independent variable.

Additionally, we analysed cost components to ascertain which contributed the highest and lowest proportions of total average costs across PHC providers. Cost component definitions are provided in **Table S3 (Supplementary Material)**.

We inspected the dataset for the level of missing or duplicated data to determine whether any recoding or adjustments to the statistical analysis was necessary.

Depending on whether cost-per-person followed a normal or non-normal distribution, we used the Mann-Whitney U-test or Student’s t-test to assess if the differences in costs were statistically different across types of locality (rural vs. urban) and ownership (private vs. public) (41).

We used univariate linear regression to identify which explanatory variables were strongly correlated with cost-per-person to guide the development of the multivariable linear regression (MLR) models (42).

We developed two MLR models, each focusing on one of the primary explanatory variables – locality and ownership type. We adjusted both models for potential confounding by incorporating other relevant explanatory variables. Following univariate linear regression, we used a forward selection (FS) approach to develop the MLR model, whereby additional explanatory variables (aside from the primary explanatory variables i.e., locality or type of ownership) are iteratively included in the model, until there are no variables left that make a significant effect (i.e., p<0.20) on the response variable (42–45). We chose FS because it includes all variables for model inclusion, and can develop more parsimonious (simpler) models which are less susceptible to multicollinearity and overfitting, therefore making them more generalisable to new data (44). Additionally, we chose a p-value entry-to-model threshold of 0.20 due to evidence from academic literature (44,46,47)

During model development, we excluded the “oblast” (i.e. county) variable in favour of the “region” variable, as the latter captured similar information with fewer parameters, enhancing parsimony.

The equation for multivariable linear regression is (42):

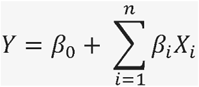

where:

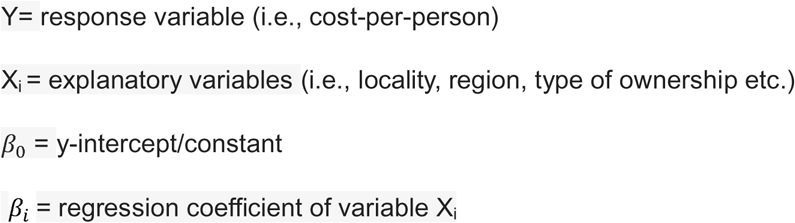

We performed a sensitivity analysis to assess whether a generalized-linear model (GLM) with a gamma distribution provided a better model fit for the final model specification compared to the MLR model. We assessed model goodness-of-fit using the Akaike Information Criterion (AIC) and Bayes Information Criterion (BIC). MLR assumes that residuals are normally distributed (48,49), which may not be appropriate for cost data which often is skewed (50–53). Therefore, we chose the GLM due to its flexibility in handling non-normally distributed data. (50). We also specifically chose a gamma distribution as it is frequently applied in health economics where costs often exhibit positive skewness (50–53).

## Results

### Descriptive statistics

**Table 2** shows the frequency distribution of the explanatory variables, cross-tabulated against PHC provider locality and ownership type. Public-urban PHC providers were the most common (56%), while private-rural PHC providers were the least common (5%). The majority of rural PHC providers fell into the lowest categories for several explanatory variables, such as average numbers of registered persons, doctors and employees. Geographically, rural providers were concentrated in the west (63%), particularly in the Rivne and Chernivtsi oblasts (16% for both). In contrast, urban providers exhibited a more even geographical distribution across Ukraine, with the central region (33%) and Odessa oblast (17%) having the highest proportions. Similarly, many private providers fell into the lowest categories for several explanatory variables, including number of service location points, as well as average numbers of registered persons and employees. Geographically, private providers were concentrated in the south (40%), with the highest percentage in Odessa (37%). Conversely, most public providers were in the west (40%), particularly in the Vinnytsia oblast (13%).

**Table 2:**
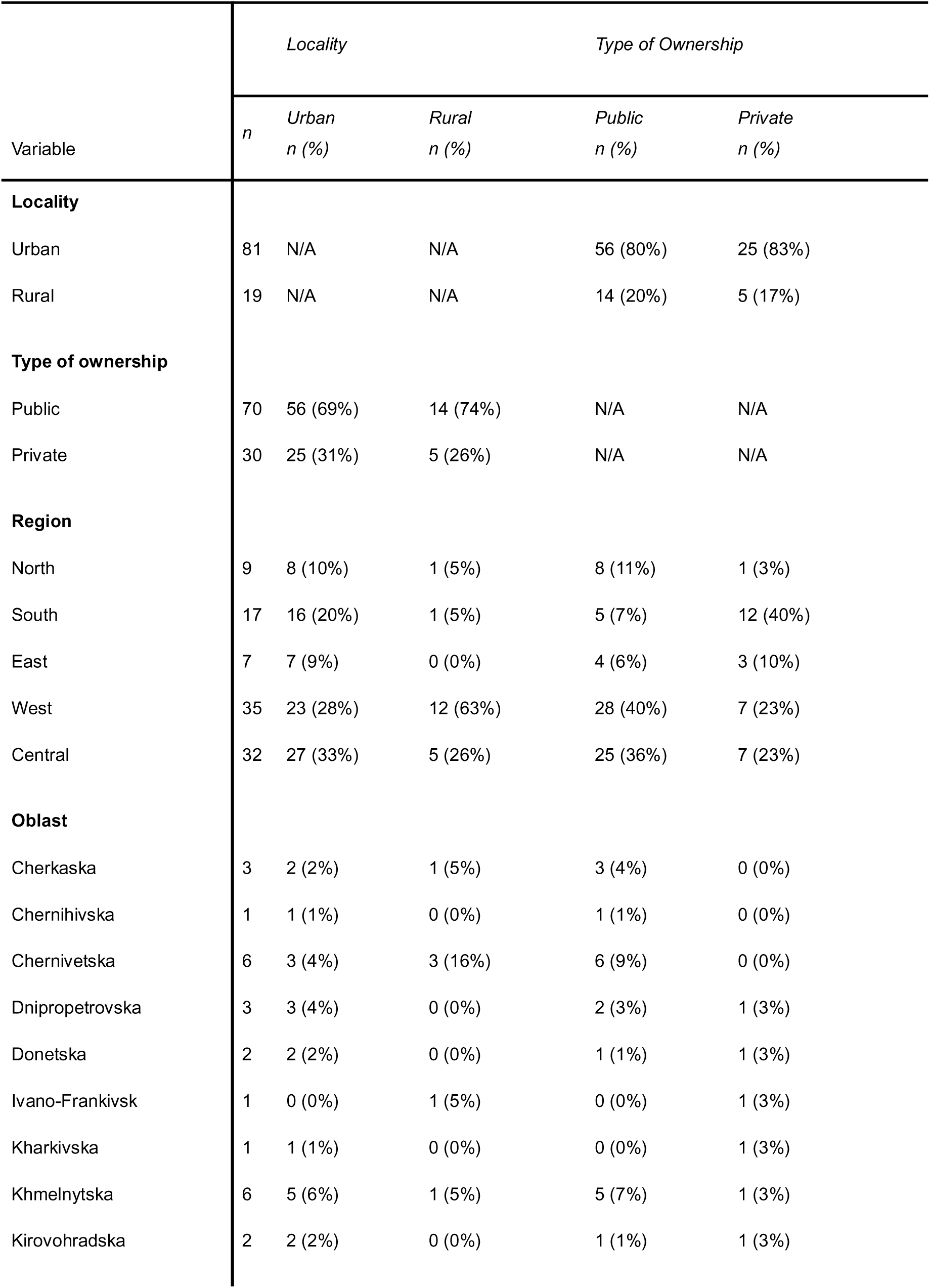
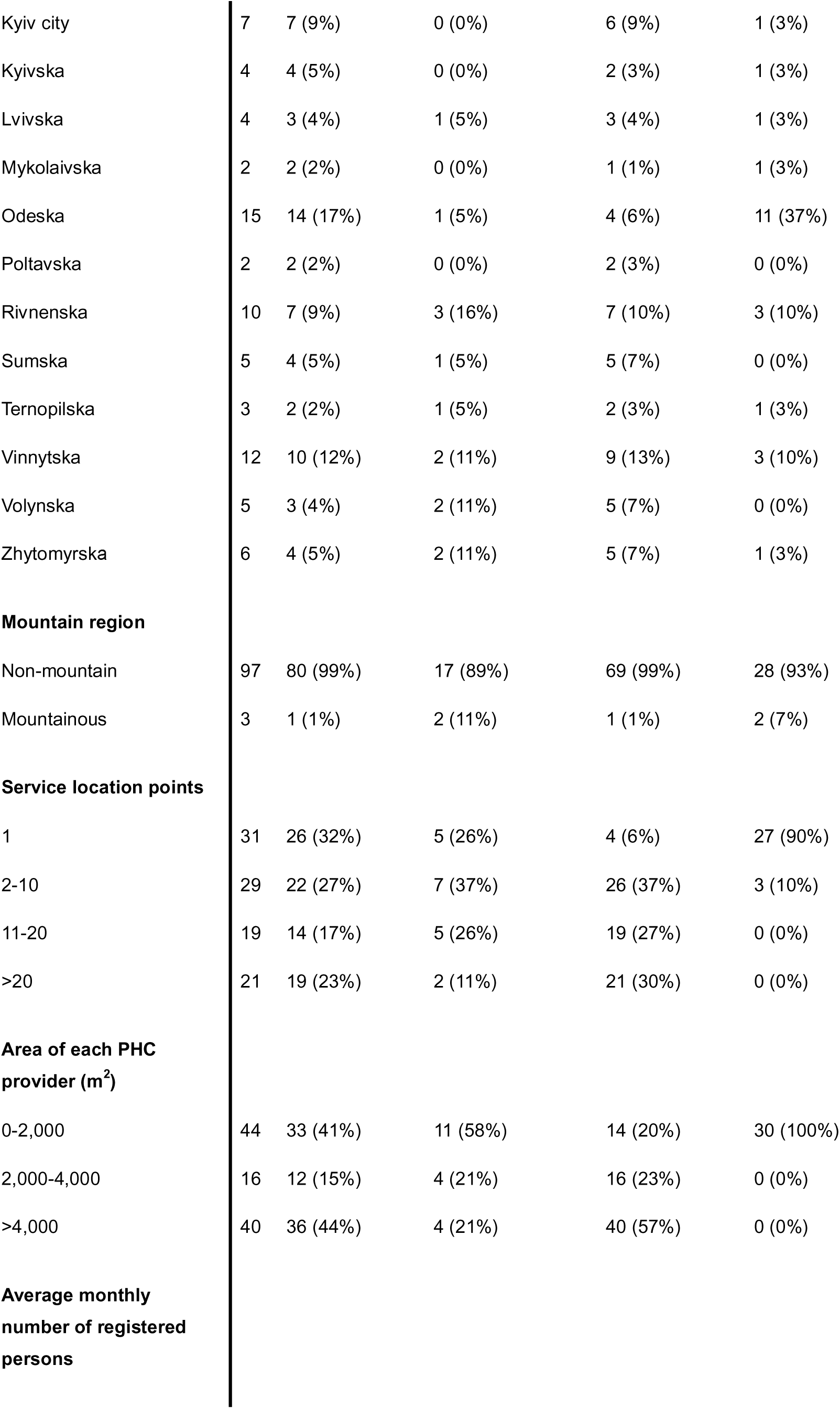
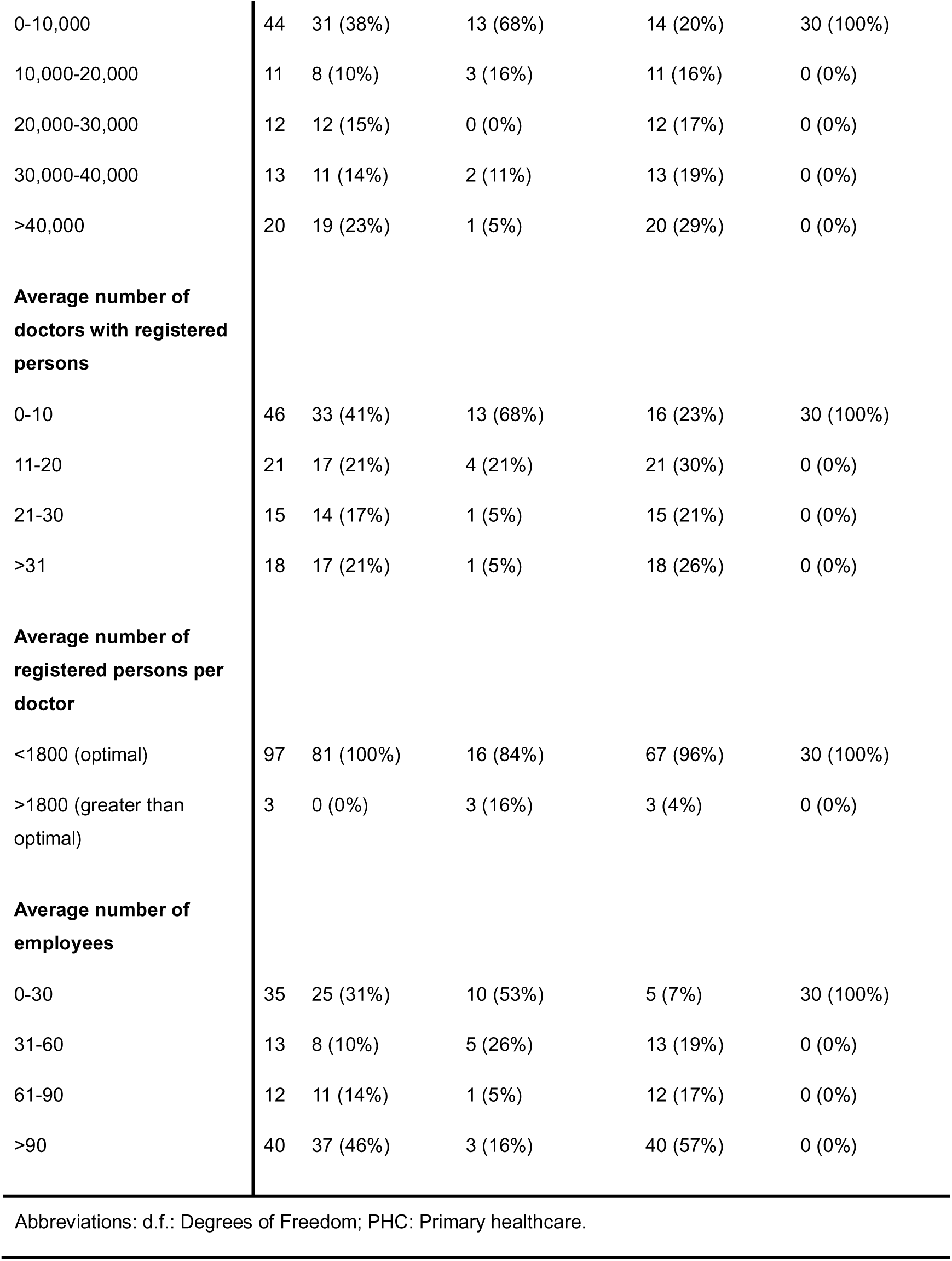
Distribution of explanatory variables, cross-tabulated against primary healthcare provider locality and ownership (n=100).

Exploratory analyses revealed no dataset errors, such as incorrect cost component totalling. However, 7 PHC providers had missing values in the salary cost component. The histogram, Q-Q plot (**Figure S1** and **S2**, **Supplementary Material**) and Shapiro-Wilk test (p<0.001) indicated that cost-per-person followed a non-normal distribution.

**Table 3** presents the summary statistics of the explanatory variables, along with the mean (standard deviation) of cost-per-person for each category. The mean (standard deviation) cost-per-person across the 100 PHC providers was 45.46 (18.46) USD. The median(interquartile-range) for cost-per-person was 50.30 (35.51-54.48) USD. Rural PHC providers had a higher mean cost-per-person than urban providers (54.58 USD vs. 43.32 USD). Public PHC providers had a higher mean cost-per-person than private providers (54.90 USD vs. 23.43 USD). Mountainous PHC providers had a higher mean cost-per-person than their non-mountainous counterparts (56.69 USD vs. 45.11 USD). Northern PHC providers had the highest mean cost-per-person among all NHSU regions (61.87 USD). The Sumska oblast reported the highest mean cost-per-person among all oblasts (72.66 USD).

**Table 3:** Mean and standard deviation of cost-per-person of primary healthcare providers across the explanatory variables.

| Variable | Cost-per-person |  |
| --- | --- | --- |
|  | *n* | Mean (standard deviation), USD |
| **Locality** |  |  |
| Urban | 81 | 43.32 (16.97) |
| Rural | 19 | 54.58 (22.01) |
| **Type of ownership** |  |  |
| Public | 70 | 54.90 (10.69) |
| Private | 30 | 23.43 (13.19) |
| **Region** |  |  |
| North | 9 | 61.87 (20.04) |
| South | 17 | 26.97 (18.01) |
| East | 7 | 37.60 (19.84) |
| West | 35 | 49.58 (13.09) |
| Central | 32 | 47.87 (15.74) |
| **Oblast** |  |  |
| Cherkaska | 3 | 50.47 (4.72) |
| Chernihivska | 1 | 51.73 ( - ) |
| Chernivetska | 6 | 49.60 (7.83) |
| Dnipropetrovska | 3 | 40.20 (23.26) |
| Donetska | 2 | 32.62 (29.92) |
| Ivano-Frankivsk | 1 | 65.01 ( - ) |
| Kharkivska | 1 | 26.58 ( - ) |
| Khmelnyska | 6 | 57.24 (14.15) |
| Kirovohradska | 2 | 38.68 (17.70) |
| Kyiv city | 7 | 46.78 (13.14) |
| Kyivska | 4 | 38.20 (22.29) |
| Lvivska | 4 | 51.72 (12.08) |
| Mykolaivska | 2 | 29.15 (22.93) |
| Odeska | 15 | 26.68 (18.23) |
| Poltavska | 2 | 57.63 (1.14) |
| Rivnenska | 10 | 44.91 (17.39) |
| Sumska | 5 | 72.66 (17.65) |
| Ternopil'ska | 3 | 38.16 (6.51) |
| Vinnytska | 12 | 47.18 (10.65) |
| Volyn'ska | 5 | 51.78 (3.00) |
| Zhytomyr'ska | 6 | 55.63 (24.87) |
| **Mountain region** |  |  |
| Non-mountainous | 97 | 45.11 (18.52) |
| Mountainous | 3 | 56.69 (14.02) |
| **Service location points** |  |  |
| 1 | 31 | 26.43 (15.86) |
| 2-10 | 29 | 51.67 (13.40) |
| 11-20 | 19 | 58.20 (13.21) |
| >20 | 21 | 53.42 (7.91) |
| **Area of each PHC provider (m<sup>2</sup>)** |  |  |
| 0-2,000 | 44 | 35.30 (23.01) |
| 2,000-4,000 | 16 | 55.04 (8.20) |
| >4,000 | 40 | 52.79 (6.77) |
| **Average monthly number of registered persons** |  |  |
| 0-10,000 | 44 | 36.42 (24.07) |
| 10,000-20,000 | 11 | 54.42 (6.47) |
| 20,000-30,000 | 12 | 54.68 (9.11) |
| 30,000-40,000 | 13 | 51.88 (6.49) |
| >40,000 | 20 | 50.69 (4.48) |
| **Average number of doctors with registered persons** |  |  |
| 0-10 | 46 | 37.05 (23.73) |
| 11-20 | 21 | 53.56 (5.83) |
| 21-30 | 15 | 53.52 (9.31) |
| >31 | 18 | 50.75 (4.73) |
| **Average number of registered persons per doctor** |  |  |
| <1800 (optimal) | 97 | 44.96 (18.43) |
| >1800 (greater than optimal) | 3 | 61.37 (12.95) |
| **Average number of employees** |  |  |
| 0-30 | 35 | 30.38 (22.79) |
| 31-60 | 13 | 59.62 (10.14) |
| 61-90 | 12 | 50.16 (4.05) |
| >90 | 40 | 52.64 (6.71) |

Abbreviations: PHC: Primary healthcare

A trend was observed where the mean cost-per-person initially increased as the number of service location points, average monthly registered persons, number of doctors, employees, and PHC provider area grew. However, beyond certain thresholds (service location points: >20; average registered persons: >20,000; doctors: >20; Employees: >60; Area: > 4,000m^2^), further increases in these variables led to a decrease in the mean cost-per-person. This trend was mirrored when the median cost-per-person was analysed as the cost-per-person data followed a non-normal distribution which is provided in **Table S4 (Supplementary Material)**

The total average costs of each cost component across the 100 PHC providers (**Figure 2**) shows that the largest contributors to total average costs were labour (81%), pharmaceuticals and biologics (7%) and medical equipment, services and related costs (3%). Conversely, medical device depreciation, software depreciation, and transport depreciation represented the smallest proportions of total average costs.

**Figure 2:**
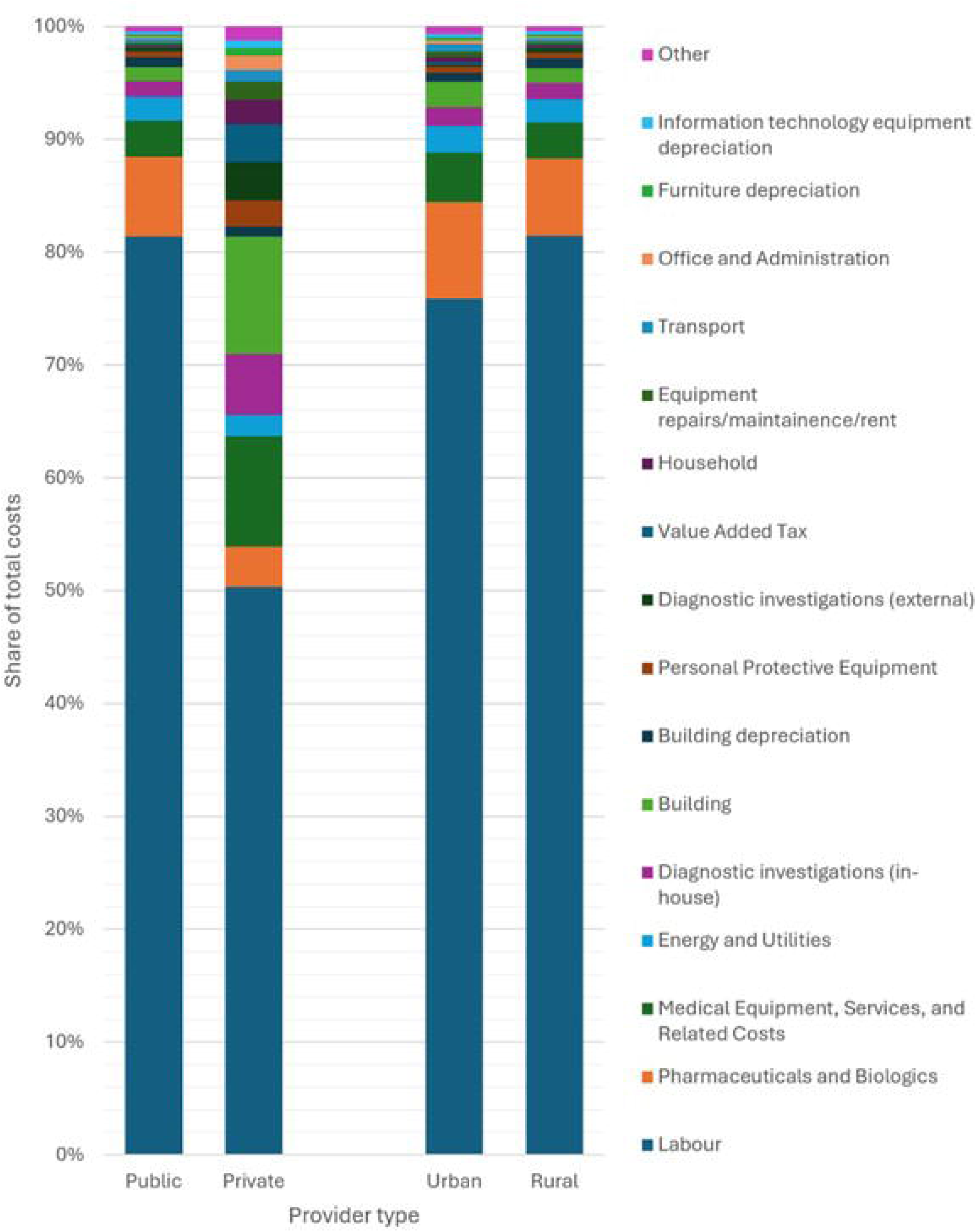
Main cost components as a percentage of total average cost. Note – Other includes costs of: Medical device depreciation, Software depreciation, Transport depreciation, Non-medical equipment, Food, Information technology, Social Security, Household equipment depreciation, Value Added Tax, Non-medical equipment depreciation, Office and Administration, Disinfectants, Furniture depreciation, Household, Information technology equipment depreciation, Diagnostic investigations (outsourced). **Alt text**: Bar chart outlining the percentage of the total average cost by each cost component.

**Figure 3** reports the percentage distribution of the top 10 combined cost components, broken down by locality (urban vs. rural) and ownership (private vs. public). Across both figures, labour, pharmaceuticals and biologics, and medical equipment, services and related costs consistently ranked as the top cost contributors.

**Figure 3:**
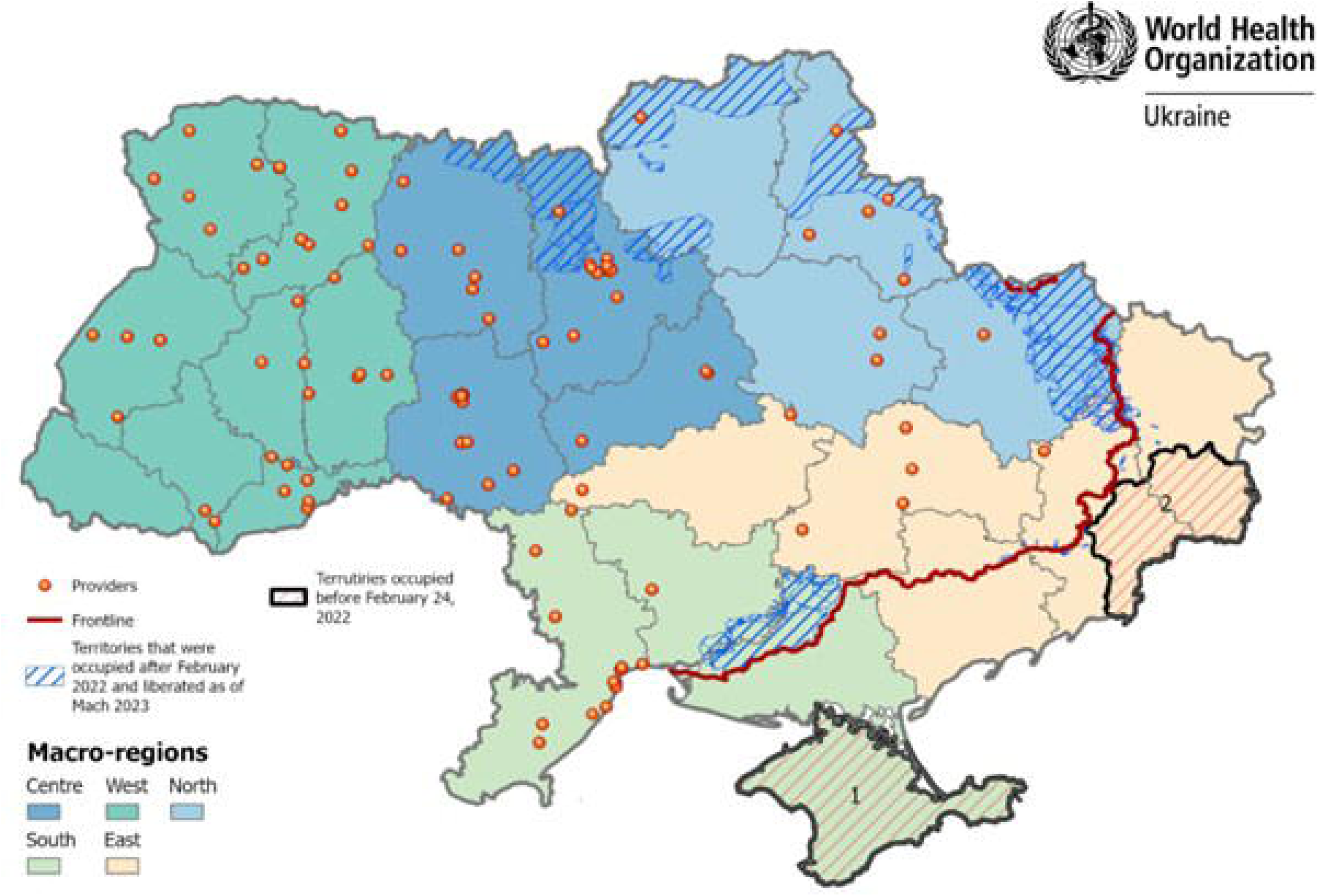
Distribution of top 10 cost components as a percentage of total average cost for rural, urban, public and private primary healthcare providers Note – Other includes costs of: Non-medical equipment depreciation, Household equipment, depreciation, Social Security, Disinfectants, Information technology, Food, Non-medical equipment, Transport depreciation, Software depreciation, Medical device depreciation. Alt text: Bar chart outlining the distribution of top 10 cost components as a percentage of total average cost for rural, urban, public and private primary healthcare providers.

However, the composition and ranking of the top cost components differed when stratified by locality and ownership (**Figure 3**). Rural and urban providers had similar top 10 cost components, with one key difference – rural providers included transport in their top 10, while urban providers included diagnostic investigations. Private and public providers differed in their top cost components. Private providers had value-added-tax, diagnostic investigations and household expenses in their top 10. In contrast, public providers had equipment repairs/maintenance/rent, transport and building depreciation.

### Cost comparison analysis

Given the non-normal distribution of cost-per-person, the Mann-Whitney U-test showed quite strong evidence of a potential statistical difference in cost-per-person between rural and urban PHC providers (p=0.0324), leading to rejection of the null hypothesis of no statistical difference. Similarly, there was very strong evidence of a statistical difference in cost-per-person between private and public PHC providers (p<0.001), also leading to rejection of the null hypothesis.

### Univariate Linear Regression

Univariate linear regression results, as shown in **Table S5 (Supplementary Material)**, show that there is strong evidence that southern and eastern PHC providers, as well as, private ownership had the highest cost-per-person with beta-coefficients of −34.90 USD (95%-CI: −47.98, −21.82, p<0.001), −24.27 USD (95%-CI: −40.27, −8.28, p=0.003) and −31.47 USD (95%-CI: −36.44, −26.50, p<0.001), respectively. Overall, except for mountainous regions, most explanatory variables showed strong evidence of a statistical relationship with cost-per-person (p<0.050).

### Multivariable Linear Regression

Using FS, two MLR models were developed to examine the relationship between locality and ownership type with cost-per-person, adjusting for confounding. Notably, FS yielded similar models for both locality and ownership type (**Table 4**), as each model included the same set of confounding variables (excluding the primary variable under investigation i.e., locality and ownership type). Both models also explained 77% of the variance in cost-per-person (R^2^=0.77, adj-R^2^=0.75, F(9, 90)=34.22, p<0.001 for both). **Table 4** shows quite strong evidence that rural PHC providers had a higher cost-per-person of 6.70 USD (95%-CI: 1.54, 11.85) compared to urban providers, after adjusting for confounding by ownership type, average registered persons per doctor, mountainous regions, average number of doctors with registered persons, and region variables (p=0.011). **Table 4** also highlights that there is strong evidence that private PHC had a lower cost-per-person of 36.15 USD (95%-CI: −41.82, −30.48) than public providers, after adjusting for confounding by mountainous regions, average registered persons per doctor, locality, average number of doctors with registered persons, and region variables (p<0.001).

**Table 4:**
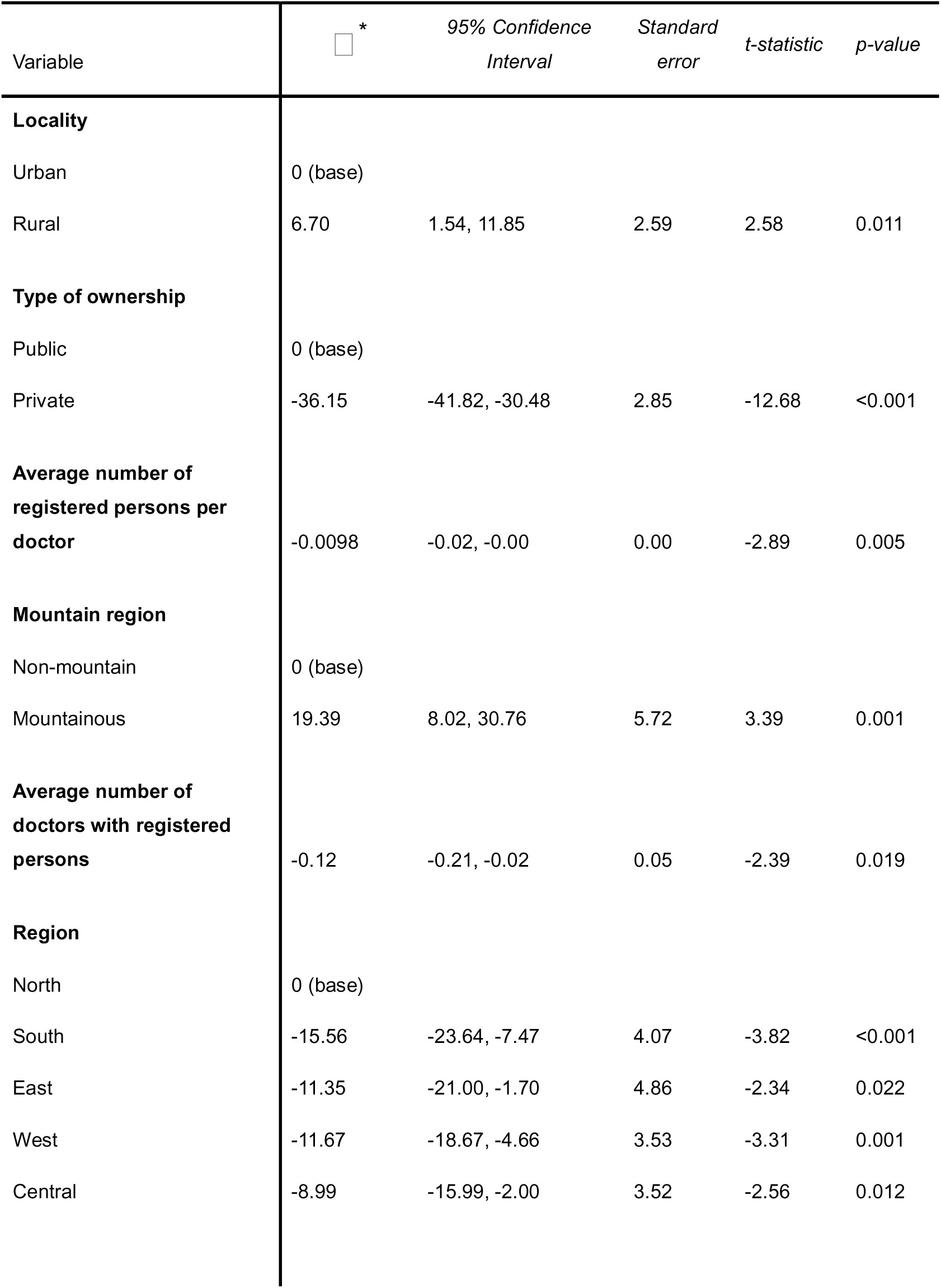
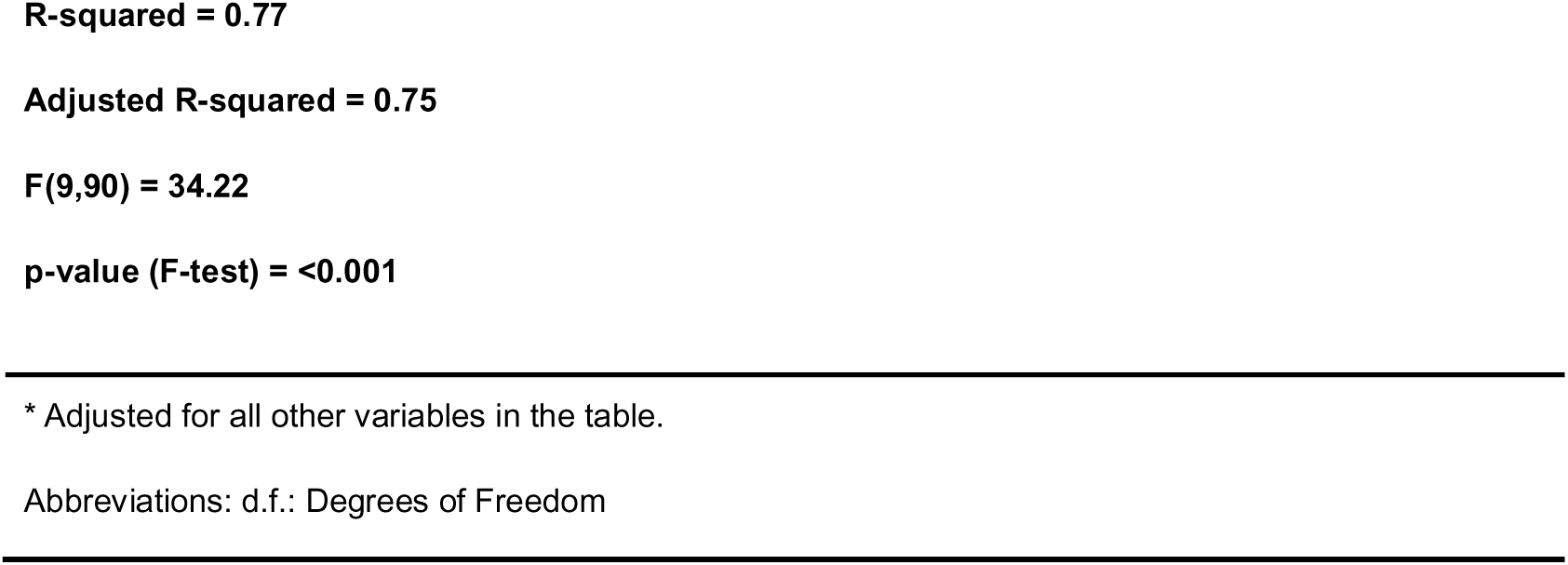
Multivariable linear regression model summary examining the association between the primary explanatory variables (primary healthcare provider locality and ownership type) and cost-per-person after adjustment for confounding explanatory variables (n=100).

| Variable | $\beta$ * | 95% Confidence Interval | Standard error | t-statistic | p-value |
| --- | --- | --- | --- | --- | --- |
| **Locality** |  |  |  |  |  |
| Urban | 0 (base) |  |  |  |  |
| Rural | 6.70 | 1.54, 11.85 | 2.59 | 2.58 | 0.011 |
| **Type of ownership** |  |  |  |  |  |
| Public | 0 (base) |  |  |  |  |
| Private | -36.15 | -41.82, -30.48 | 2.85 | -12.68 | <0.001 |
| **Average number of registered persons per doctor** |  |  |  |  |  |
|  | -0.0098 | -0.02, -0.00 | 0.00 | -2.89 | 0.005 |
| **Mountain region** |  |  |  |  |  |
| Non-mountain | 0 (base) |  |  |  |  |
| Mountainous | 19.39 | 8.02, 30.76 | 5.72 | 3.39 | 0.001 |
| **Average number of doctors with registered persons** |  |  |  |  |  |
|  | -0.12 | -0.21, -0.02 | 0.05 | -2.39 | 0.019 |
| **Region** |  |  |  |  |  |
| North | 0 (base) |  |  |  |  |
| South | -15.56 | -23.64, -7.47 | 4.07 | -3.82 | <0.001 |
| East | -11.35 | -21.00, -1.70 | 4.86 | -2.34 | 0.022 |
| West | -11.67 | -18.67, -4.66 | 3.53 | -3.31 | 0.001 |
| Central | -8.99 | -15.99, -2.00 | 3.52 | -2.56 | 0.012 |

### Sensitivity Analysis

Given that FS yielded models with the same set of confounding variables, the final model specification for cost-per-person included locality, ownership type, average number of registered persons per doctor, mountain region, average number of doctors with registered persons, and region variables. Sensitivity analysis revealed that the GLM with the final model specification (**Table S6**, **Supplementary Material)** and gamma distribution had a lower goodness-of-fit (AIC=966.15, BIC=992.20) compared to the MLR model with the final model specification (AIC=736.38, BIC=762.43).

## Discussion

We observed that the largest contributors to total average costs of PHCs in Ukraine were labour, which accounts for 81%, followed by pharmaceuticals and biologics at 7%. This aligned with other studies indicating that major drivers in PHC delivery costs are labour and medicines (54,55). However, cost components varied in content and order when stratified by locality and ownership type. The average (standard deviation) cost-per-person across the providers was 45.46 (18.46) USD which is significantly higher compared to the paid NHSU base capitation of 651.60 UAH (23.89 USD) in 2021 (8).

From MLR, we found strong evidence that rural PHC providers had a higher cost-per-person of 6.70 USD (95%-CI: 1.54, 11.85) compared to their urban counterparts, after adjusting for the confounding (p=0.011). We s also found very strong evidence that private PHC providers had a lower cost-per-person of 36.15 USD (95%-CI: 41.82, 30.48) compared to their public counterparts, after adjusting for confounding (p<0.001).

### Divide by provider characteristics

We observed a trend where the mean (and median) cost-per-person initially increased as the number of service location points, average monthly registered persons, number of doctors, employees, and PHC provider area grew. This observation may be partly explained by these Ukrainian PHC providers benefiting from economies of scale – where long-run average costs increase as output increases (56,57). Economies of scale arize when fixed costs are spread over more registered persons, thus reducing the cost-per-person (14,22,25,56,57). However, beyond a certain threshold, further increases in these variables led to an increase in the mean (and median) cost-per-person. Our findings indicated that providers with more than 20,000 registered patients, 20 doctors, and 40 total employees had higher per-registered person costs. Similarly, providers with more than 10 service location points also incurred more costs per registered person. Additionally, providers operating in smaller facilities (measured in total square meters) also had lower per-person costs. This insight may be valuable for purchasers in developing financial incentives — such as defining an optimal facility size for cost coverage — to balance cost efficiency with maintaining optimal service standards.

### Rural-Urban divide

The results suggested that rural Ukrainian PHC providers faced a higher cost-per-person than their urban counterparts. This result was consistent with similar findings from other global studies (14,20–23,25,26,29,30). Several factors could have contributed to the rural-urban cost difference. Lower population densities in rural areas could have led to longer travel distances for both providers and patients, making healthcare delivery more time-consuming and expensive (14,20,21,25,58–64). This issue may have been exacerbated by poor public transport (14,22,59,60,62,63,65) and infrastructure – a particular issue in rural Ukraine (14,60,63). These factors may have contributed to transport being a larger cost component for rural providers. Additionally, rural areas often have greater health needs than urban areas, partly due to elderly populations and higher rates of adverse health-behaviours like smoking and poor diets, which may stem from rurality being associated with lower educational attainment and health literacy (14,20,21,23,25,62–64). This may have collectively contributed to greater healthcare demand by rural Ukrainian residents, which resulted in more frequent PHC visits (66), and therefore increased provider costs compared to urban areas.

Similarly, rural areas in Ukraine and globally, are known to have poorer facilities and equipment (14,20,23,25,26,58,64,67). A Ukrainian rural healthcare survey found that ‘lack of required services’ and ‘redirection to other providers’ were particular challenges to healthcare access (60), which were further intensified by rural staff shortages (8,68). The WHO health labour market analysis indicated that, while approximately 30% of Ukraine’s population lives in rural communities, only 17% of PHC doctors and 7% of PHC nurses serve these areas, highlighting a significant workforce gap (68). Additionally, barriers such as increased travel time (and thus costs), poor infrastructure, and inadequate services may act to deter rural residents from seeking timely PHC care. This delayed presentation may cause their condition(s) to worsen, and become more complex (and perhaps costly) to treat (64,69,70). Moreover, these barriers may have encouraged rural residents to register with better-equipped urban PHC providers for better care. According to January 2024 data, 43% of PHC services locations were in small villages with a population up to 5,000 persons. Among the physicians working there, 55% had fewer than 1,500 registered persons (10). Urbanisation may have left rural PHC providers with fewer patients to cover fixed costs, resulting in a higher cost-per-person (14,22,25,56) as well as raising concerns about financial sustainability and limited provider interest in these areas. Therefore, many rural providers are owned by local governments.

The capitation rate is adjusted with a coefficient of 1.2 for 3% of PHC providers providing services in mountainous areas, but there are currently no funding adjustments for providers serving rural areas. Because of this, local governments in rural areas often must compensate for the extra costs to ensure access to PHC. Local governments fund healthcare from their own revenues, including utility and capital costs of the facilities, infrastructure investments, salary top-ups, or compensations like accommodation for healthcare professionals etc. These additional expenditures are substantial, but they are also highly variable across regions (7). To ensure equitable funding, independent of local government financial capability and ownership, capitation adjustments could be applied to rural providers as a financial incentive. International experience shows that capitation adjustments are extensively used when there is proof of significant cost variations (3). Capitation adjustment coefficients could be based on evidence from this analysis which indicated that rural providers in the sample had approximately 20% higher expenditures compared to providers in urban areas.

### Private-Public divide

Our results suggested that public PHC providers had a higher cost-per-person compared to their private counterparts. This may have been due to several factors. Private PHC providers reportedly faced lower administrative burdens (8), potentially due to employing fewer staff and utilising smaller rooms compared to their public counterparts. The smaller size of private practices may have enabled their doctors to handle administrative tasks themselves, reducing the need for additional administrative employees, in turn decreasing the cost-per-person (71). Private providers may have also benefited by outsourcing administrative functions, enabling them to focus on core clinical work (72–75), therefore, avoiding the potentially expensive costs associated with scaling in-house capacity (73,76,77). Private providers could have also established their own staffing structures, independent of the standards adhered to by public providers. For instance, public providers are expected to meet specific staffing standards, ensure facilities comply with state sanitary guidelines, adhere to opening hours set by local governments (which own public PHC facilities), and fulfil more extensive reporting requirements to NHSU (see **Table 5**). FOPs in the cost study sample employed on average, only half of the number of staff compared with public providers, and some did not employ a nurse at all (8). However, cutting costs by failing to meet staffing and other standards may have compromised care quality, prioritising savings over patient well-being with an objective of profit maximization. Similar trends have been observed in the UK, where private PHC providers replaced doctors with less qualified staff to maximize profits (78).

**Table 5:**
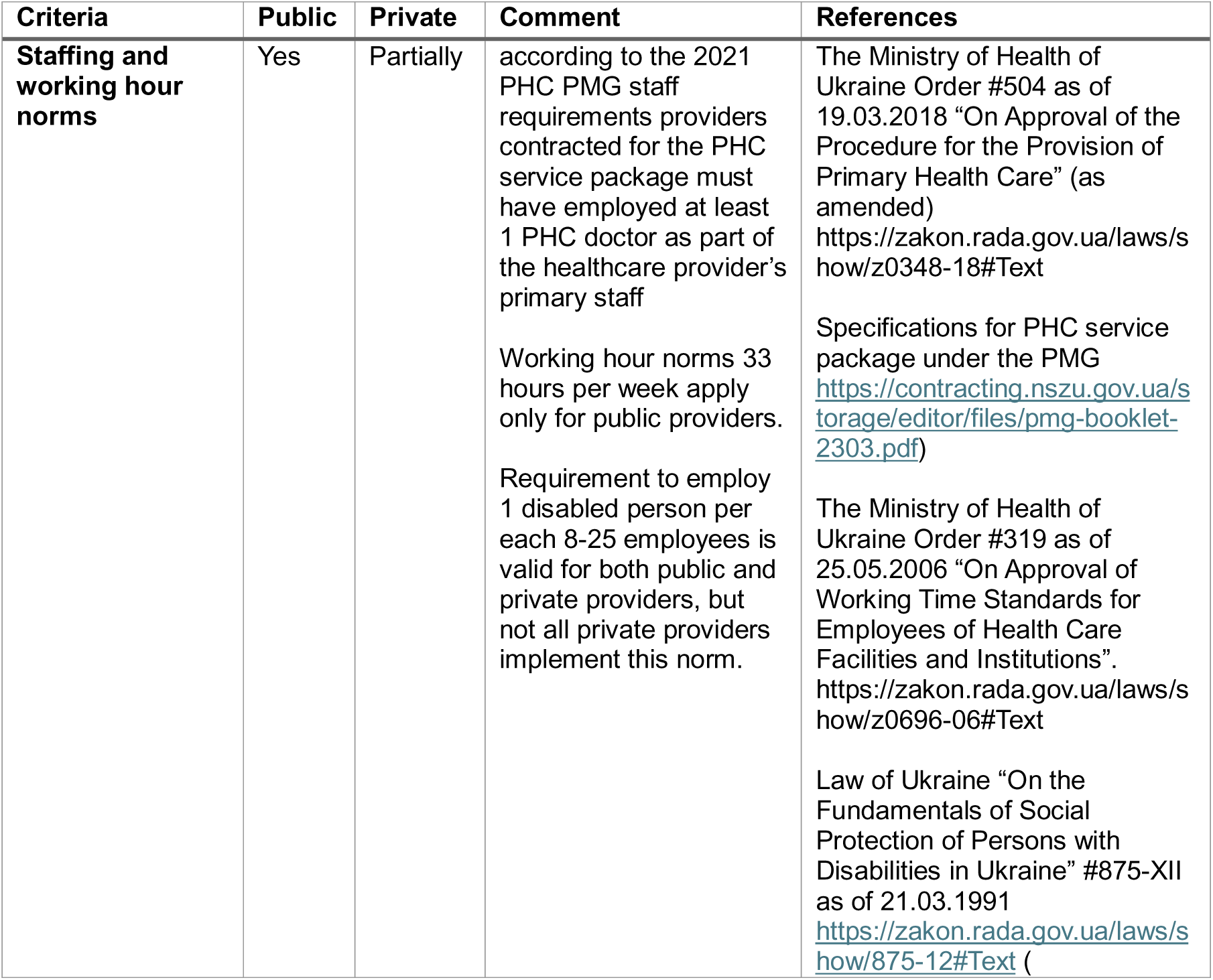
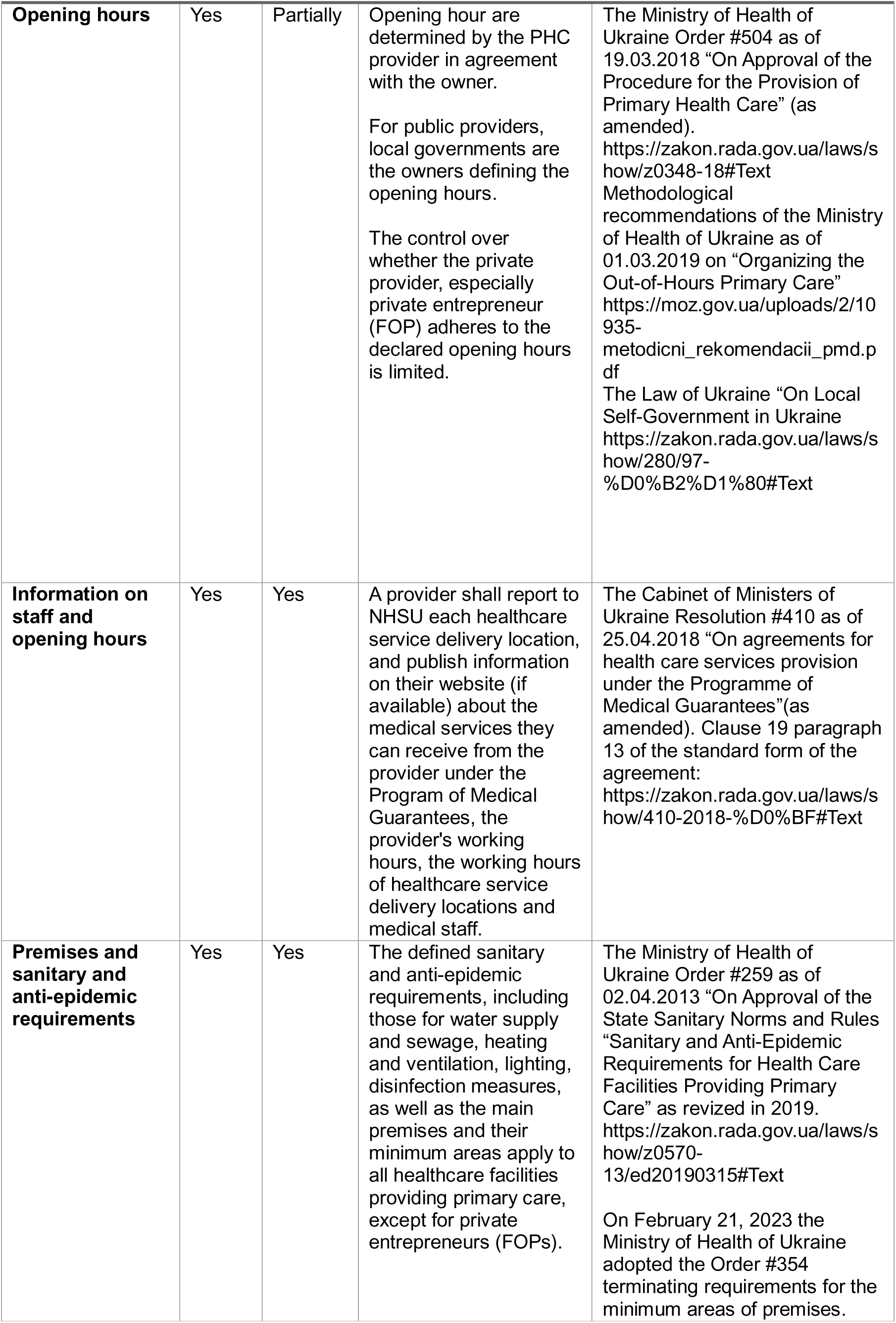
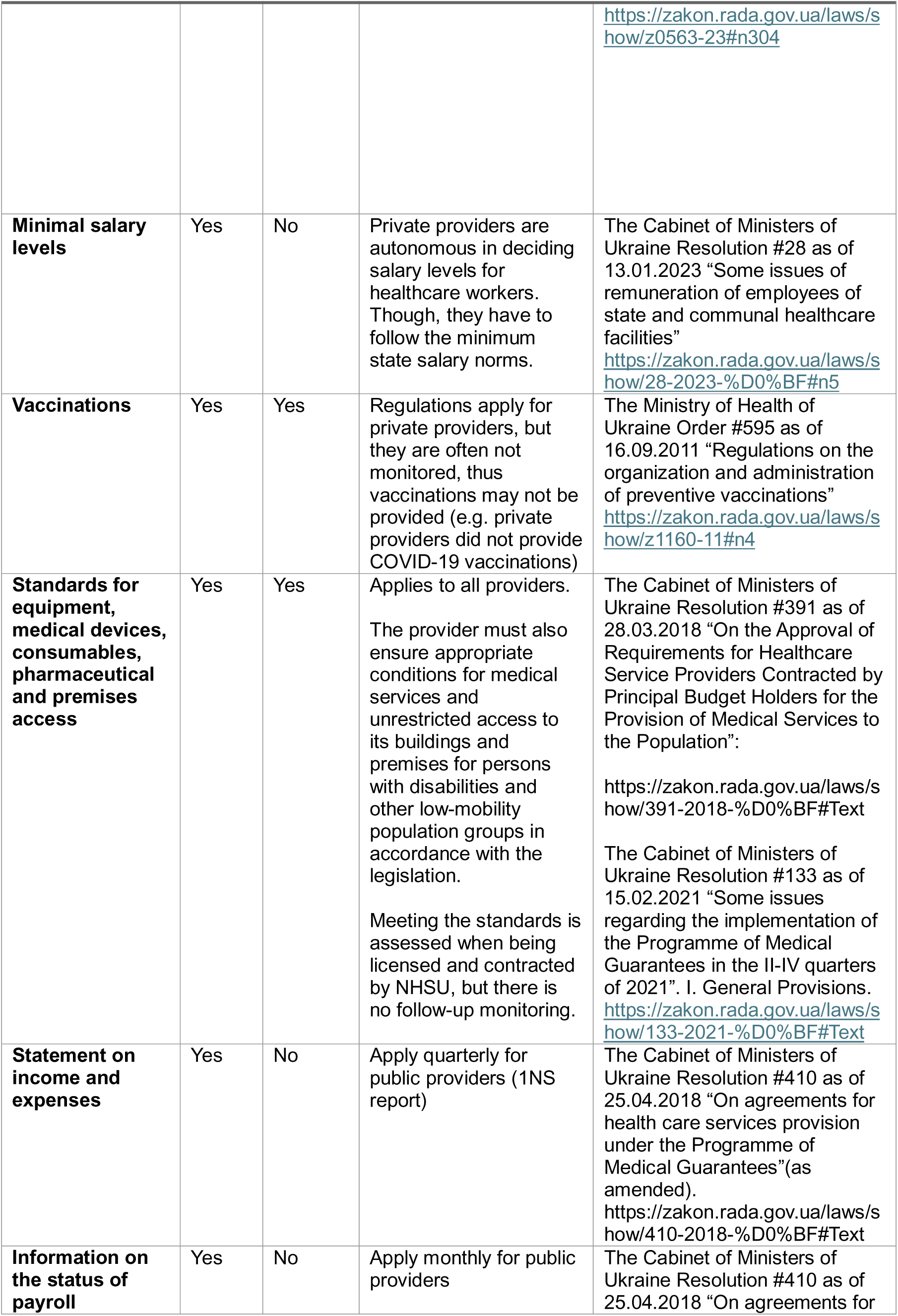
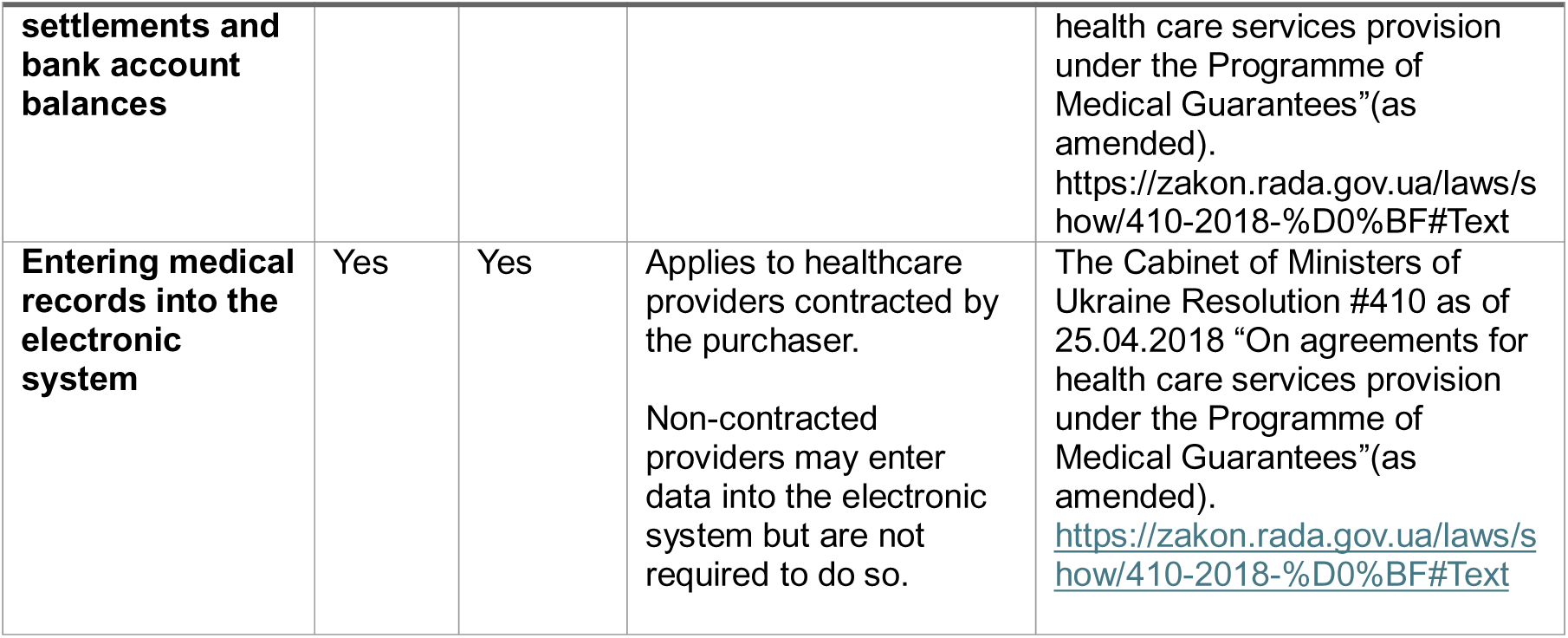
Difference in requirements for public and private providers.

Our analysis revealed that the cost components constituting a greater proportion of the total average cost differed between private and public providers. For example, private providers spent more on building and household expenses, likely due to not having access to local government funds compared to public providers (8). Our results also showed that private PHC providers spent a greater proportion of their total average cost on external diagnostic tests compared to public PHC providers which spent a greater proportion on in-house diagnostic tests. This could partly explain the higher cost-per-person for public providers as it may have been more expensive to provide in-house testing for several reasons. External labs might have been more specialized in handling large volumes of tests (and its associated admin), allowing greater operational efficiency (perhaps through internal specialisation), and at a lower cost compared to in-house services (72–75,79,80). Nonetheless, in Ukraine, data on lab services delivered under the PMG were not collected and therefore cost differences between public and private providers may have been explained by patients shifting towards privately paid services.

Additionally, private providers have been reported to engage in cream-skimming, by selecting younger populations for their patient lists (8), who may have been less likely to require extensive PHC services, thereby reducing their cost-per-person (81–84). This may have been driven by a strategic decision by private providers to concentrate on urban areas rather than serving low population-density regions or areas with a higher proportion of elderly residents whereby service delivery costs are higher and revenues lower. This trend aligned with academic literature which showed that urban areas typically have more private healthcare providers compared to rural areas, including in Ukraine (8,17,25,27,85–87).

Overall, our findings suggested that lower costs-per-person may have been better explained by private providers being more interested in cutting costs for profit maximization, rather than providing “better value for money” compared to public providers. The standards defined for PHC service delivery in Ukraine are often not enforced on private providers, allowing them to compromise in healthcare service quality and access. Although the academic literature on the relationship between healthcare costs and provider ownership is varied, highlighting the influence of country-and institution-specific contexts (15–19,24,27,28), our findings indicated that Ukraine could increase “value for money” spent publicly on PHC by strengthening regulatory oversight over private providers. This would help to protect the interests of patients and balance the opportunistic behaviour for profit maximization by private providers.

### Strengths and Limitations

Our study had several strengths. We used provider-level cost data, filling a significant literature gap of healthcare costing studies in Ukraine, as well as across Eastern Europe. While there is limited costing research in Eastern Europe, similar studies (including from other low-middle income countries) aligned with this study’s findings (14,20–23,25,26,29,30). The methodology of this study is comprehensive and grounded in academic literature, which enhances the study’s internal and external validity, result generalisability, while also providing a transparent standardized methodology to enhance replicability. PHC providers were screened based on a predefined selection criteria and their willingness to participate in the data collection. This convenience sampling helped ensure a representative sample of the PHC provider distribution across Ukraine (12), strengthening the study’s validity. Also, 100 PHC providers were selected in the PHC costing study to achieve a theoretical sample error of 10%, with a 95%-CI (12). Additionally, with less than 10% missing data, no recoding or alterations were needed for the statistical analysis (88). Sensitivity analysis further supports the robustness of the study, showing that the MLR models had the better goodness-of-fit (lower AIC and BIC values) for the final model specification compared to a GLM using a gamma distribution.

However, despite these strengths, this study also had limitations. This retrospective cross-sectional study relied on 2021 costing data, limiting the ability to conduct a longitudinal analysis of historic Ukrainian PHC costing trends. Additionally, it did not account for the impact of Russia’s invasion of Ukraine, which affects the relevance of the findings for current policy development. The invasion has strained the Ukrainian PHC system, particularly with healthcare staffing shortages, and increasing prevalence of certain health conditions (8,89) – both of which could increase healthcare costs. The retrospective nature of data collection may have introduced recall bias and non-differential misclassification measurement errors, limiting internal validity. Additionally, cost-per-person was the only response variable analysed in this study, meaning no specific inferences about efficiency can be reliably made. The study also excluded the Kherson, Luhansk Zakarpattia and Zaporizhia oblasts (due to inability for providers to share data as a consequence of the Russian full scale invasion, potentially affecting its generalisability. Additionally, this study focused on the PHC provider perspective, missing insights into patient out-of-pocket spending on PHC including medicines, which is a critical issue in Ukraine (6,8,67,90). The limited costing literature in Eastern Europe also hinders comparative analysis.

## Conclusion

Strong PHC is the foundation of a resilient healthcare system, improving population health outcomes while enhancing efficiency and reducing overall costs. Despite major reforms to improve financing equity, further reform is needed for the Ukrainian PHC system. The public budget allocated to health remains challenged by the ongoing hostile invasion by the Russian Federation (8,12,67,91), which has created the need for the NHSU to ensure that the limited budget is spent where it provides most value.

This secondary data analysis of 2021 costing data from the Ukrainian PHC costing study found that rural and public PHC providers had a higher cost-per-person compared to their urban and private counterparts, even after adjusting for confounding. It also showed that the cost-per-person depended on different provider level characteristics. This information is valuable for purchasers in developing financial incentives to optimize service delivery costs, but also to ensure that minimum standards are met to facilitate high quality-of-care. The NHSU could initiate capitation adjustments for rural providers as a financial incentive to enable PHC service access and continuity for rural populations. Ukraine should also review its approach to enforcing regulations on PHC providers, ensuring that all providers, regardless of ownership type, adhere to standards that guarantee high-quality accessible PHC services. There is a clear need to strengthen regulatory oversight of private providers to ensure better “value for money” and protect patient interests, while balancing their desire for profit-maximization.

To further facilitate PHC financing reform in Ukraine, additional and continuous cost data collection and analysis should be encouraged to explore cost drivers and validate our findings. Ukraine would benefit from establishing a national PHC costing database which would allow enhanced data quality, support longitudinal analysis, and enable evidence-based policy adjustments, particularly amid the challenges of the ongoing war. By adopting these recommendations, Ukraine can build a more resilient, data-driven PHC financing system to improve health outcomes and support recovery efforts.

## Abbreviations

95%-CI: 95% Confidence Interval
AIC: Akaike Information Criterion
BIC: Bayesian Information Criterion
FS: Forward Selection
GHCC: Global Health Cost Consortium
GLM: Generalized Linear Model
MLR: Multivariable Linear Regression
NHSU: National Health Service of Ukraine
PHC: Primary healthcare
PMG: Program of Medical Guarantees
UAH: Ukrainian Hryvnia
USD: United States Dollar
WHO: World Health Organization

## Declarations

### Ethical Approval

Ethical approval for this analysis was granted by the LHSTM MSc Research Ethics Committee (Ref: 30475) on 29th April 2024.

### Consent for Publication

Not applicable

### Availability of data and materials

The data underpinning this study will be shared at reasonable request by the corresponding author.

## Competing interests

All authors have no conflicts of interest to declare.

## Funding

The authors gratefully acknowledge support from the Government of Canada and WHO Universal Health Coverage Partnerships for the primary healthcare cost data collection, which laid the ground for this research. The contents of this publication are the sole responsibility of the authors and do not necessarily reflect those of the Government of Canada.

## Author Contributions

ATM designed the study and the research protocol with input from KK. Data collection was done by WHO Ukraine country office. Data analysis was conducted by ATM, with support from ZS. ATM and KK drafted the manuscript, while OD, TH, AM, ZS, JH edited the manuscript. All authors contributed to interpretation of the data. All authors have seen and approved the manuscript. ATM is the guarantor and affirms that this manuscript is an honest, accurate, and transparent account of the study being reported; that no important aspects of the study have been omitted; and that any discrepancies from the study as planned (and, if relevant, registered) have been explained.

## Data Availability

The data underpinning this study will be shared at reasonable request by the corresponding author.

## Acknowledgments

The commentary uses many of the published and unpublished reports to which all members of the WHO Country Office in Ukraine have made significant contributions, working hand-in-hand with the Ministry of Health of Ukraine and the National Health Service of Ukraine.

We also thank all the Primary Healthcare providers who provided the cost data underpinning this study and all the colleagues who supported the data collection.

## Authors’ information

The authorial team consists of three males (ATM, ZS, JH) and four females (AM, KK, OD, TH) with different levels of seniority and experience across different aspects of public health. The authors include one MSc Public Health candidate and medical student (ATM), two assistant professors (AM, ZS), two healthcare financing consultants (KK, OD) and two health policy specialists (JH, TH). The authors have expertise in several areas including healthcare financing, costing studies, health economics, and health in humanitarian crises. Two of the authors are currently based in a low-and middle-income country (OD and JH), while the remaining are based in high-income countries. These range of experiences and perspectives were used in interpreting the results from this study and writing this manuscript.

## Notes

### Competing Interest Statement

The authors have declared no competing interest.

