## Supplemental Material for "Statistical Analysis of Pre-War Primary Healthcare Costs in Ukraine: Variations by Location and Ownership and Implications for Financing Reform"

Supplementary Material

### Supplemental Tables

Table S1: Inclusion criteria for the Ukrainian Primary Healthcare Costing Study.

| **Criteria** | **Description** |
| --- | --- |
| Provider type | PHC providers. |
| Number of patients per doctor | Number of patients per doctor is 900-2000. |
| Service packages provider is contracted for | PHC services package, COVID-19 vaccination package. |
| Structure of providers by regions | Structure of reference providers proportionally corresponds to the NHSU-contracted healthcare provider network as of the end of 2021, split across the five NHSU Inter-regional Departments:  Central – 20%  Eastern – 18%  Northern – 16%  Southern – 13%  Western – 33% |
| Structure of providers by ownership | Structure of reference providers proportionally corresponds to the NHSU-contracted healthcare provider network as of the end of 2021:  59% public providers  41% private providers |
| Structure of providers by locality | 22% of selected public providers were in rural areas and 78% in urban areas (i.e., cities and towns).  12% of selected private providers were in rural areas and 88% in urban areas. |
| Data collection time-period | 2021 year. |

Table S2: Methodology of the main Ukrainian Primary Healthcare Costing Study (1).

| **Methodological components** | **Description** |
| --- | --- |
| Study design | Retrospective cross-sectional study design. |
| Data source | PHC provider costing data from the 2021 financial year. |
| Data recall period | 6-10-month timespan which data was retrospectively collected/reported. |
| Costing methodology | Bottom-up costing – costing methodology where resource utilisation used to produce a service is recorded and costs assigned accordingly, to estimate costs for a system (2,3).  Normative/guideline-based – Measuring resource utilisation under defined minimum PHC standards as outlined by Ukrainian regulations.  Financial costs – the actual expenditure paid for producing goods and services, directly reflecting spending (4).  Full costs – include the sum of all costs (both direct and indirect) associated with PHC provision by providers (2).  The costing methodology was developed using Joint Learning Network resources (5). |
| Costing perspective | Provider and Purchaser perspectives enables direct evaluation of costs to providers and the NHSU through contracting. |
| Abbreviations: PHC: Primary Healthcare; NHSU: National Health Service of Ukraine. | |

Table S3: Definitions and constituent cost items for the cost components in this study.

| **Cost component** | **Notes and associated cost items** |
| --- | --- |
| Labour | Salary, Labour, Payment accruals. |
| Social security tax | N/A |
| Pharmaceuticals and biologics | Medicines and immunological preparations, blood and blood components, therapeutic nutrition. |
| Disinfectants | N/A |
| Personal Protective Equipment | Shoe covers, Skin Protective Products, Protective kits (clothing, shoe covers, shield, apron, etc.), Mask (surgical, medical, other masks), Sleeves, Protective clothing, Safety goggles, FFP2 / FFP3 respirator, Gloves (disposable, examination, other medical gloves), Apron, Isolation gown, Disposable gown, Medical cap, Shield/Screen Protector. |
| In-house diagnostic tests and investigations | Complete blood count with leukocyte count, Urinalysis, Blood Glucose, Total cholesterol, Rapid Pregnancy Tests, Rapid Troponin Tests, Human Immunodeficiency Virus (HIV) Rapid Tests, Viral Hepatitis Rapid Tests, SARS-CoV-2 Rapid Tests, Test to Detect Occult Blood in Stool, Prostate-Specific Antigen (PSA) Test, Rapid Tests for Influenza A/B, Ferritin, Coagulation, D-dimer, Procalcitonin, C-reactive protein, quantitative C-reactive Protein, Enzyme Linked Immunosorbent Assay (ELISA) blood test for viral hepatitis B, Enzyme Linked Immunosorbent Assay (ELISA) blood test for viral hepatitis C, Bacteriological Studies, pH Test, Haemoglobin Level Test, Combined Drug Detection Test, Acetone Test, IgM Examination Acute Respiratory Disease COVID, IgG Examination of Acute Respiratory Disease COVID. |
| External diagnostic tests and investigations | Complete blood count with leukocyte count, Urinalysis, Blood Glucose, Total cholesterol, Rapid Pregnancy Tests, Rapid Troponin Tests, Human Immunodeficiency Virus (HIV) Rapid Tests, Viral Hepatitis Rapid Tests, SARS-CoV-2 Rapid Tests, Test to Detect Occult Blood in Stool, Prostate-Specific Antigen (PSA) Test, Rapid Tests for Influenza AB, Ferritin, Coagulation, D-dimer, Procalcitonin, C-reactive protein, quantitative C-reactive protein, Enzyme Linked Immunosorbent Assay (ELISA) blood test for viral hepatitis B, Enzyme Linked Immunosorbent Assay (ELISA) blood test for viral hepatitis C, Bacteriological Studies, pH Test, Haemoglobin Level Test, Combined Drug Detection Test, Acetone Test, IgM Examination Acute Respiratory Disease (COVID-19), IgG Examination of Acute Respiratory Disease (COVID-19). |
| Food | N/A |
| Household | Soft inventory (costs associated with linen, mattresses, blankets, pillows etc.), Plumbing (technical means related to sewerage, heating, ventilation of premises, water, heat, and gas supply), Blinds, Curtains, Meters (gas, electricity, water, etc.), Chandelier, Lamp, Alarm system, Non-Medical Thermometers, Doors, Antenna, Gas Cylinder, Paper Towels, Toilet Paper, Liquid Soap, Hard Soap, Garbage Bags, Acetates (solution), Mobile Top-Up Cards, Cleaning Agent, Cleaning Accessories (sponges, brushes, brooms, mops, etc.), Shovel, Rake, Trowel, Materials for Repairs (paint, wallpaper, paintbrushes, putty, etc.), Spare Parts (not for Transportation), Detergent, Fire Kit (Fire Extinguisher, Axe, Shield), Water filter, Castle (Lock), Bucket, Pot, Bowl, Ladder (stepladder, etc.), Sugar Bag, Light Emitting Diode (LED) Lamps, Air Freshener, Door Closer, Bristle Coating, Door Handle, Duster Window Washer, Window Unit, Dowel, Wood Carving, Sprayer, Battery, Gutter, Plat band (Trim), Carpeting, Shockproof Double-Glazed Windows, Floor Polisher, Extension Cord, Electrical Accessories, Toilet seat, Mosquito Net, Paving Slabs, Plastic Window, Fiberglass Mesh, Tape Measure. |
| Office and administration | Medical card of the patient (inpatient/outpatient/student/child/attached sheet to the card), Certificate Forms, Medical Record Forms, Logs, Immunization Card/Immunization Forms, Sick leave certificates, Company Car Forms, Prescription forms, Ambulatory Card Separator, Power of Attorney form, Information Consent form, Ledger, Death Certificate, Hazard Warning Sign, Facade sign, Accessibility Mark, Booth, Cabinet plate, Information Plate, Tactile Tiles, Tactile Tape, Information posters, Information leaflets, Printed books, Printed magazines, Printed newspapers, E-books, Electronic Journals, Electronic newspapers, Notebook, Eraser, Hole Punch, Clamps (Binders), Notebooks, Calendar, Pushpin Button, Glue, Adhesive Tape, Stationery book, Envelope, Proofreader, Blades, Ruler, Stationary organisers, Markers, Clerical knife, Scissors, White Paper, Note Paper, Coloured paper, Folders, Seals, stamps, Pens, pencils, refills, Scotch tape, Paper clips, Stapler, Staples for staplers, Sticker, Folder, Sharpener, Files, Stamp paint, Felt-tip pens, Binder, Shiloh, Twine, Diary, Stapler-remover, Newsprint, Whatman paper. |
| Energy and Utilities | Fuel (not for transport or heating), Payment for heat supply, Payment for electricity, Natural gas payment, Payment for other energy and utilities, Payment for water supply and sewerage, fuels and lubricants for heating (pellets, firewood, coal, etc.). |
| Building | Repair of premises, Repair of elevators, Repair of the boiler room, Repair of the garage, Maintenance of buildings, structures, and premises, Maintenance of elevators, Maintenance and servicing of boiler houses, Maintenance of fire extinguishers, Maintenance/verification of meters, Calibration of household gas detectors, Gas pipeline maintenance, Gas boiler maintenance and servicing, Maintenance of electrical networks, Maintenance of water supply and sewerage networks, Maintenance of electrical installations (transformers, etc.), Premises Rent, Garages Rent, Boiler house rent. |
| Building depreciation | N/A |
| Value Added Tax | N/A |
| Repairs/Maintenance/Rent (not related to premises) | Repair of medical equipment, Repair of office equipment (including computers), Repair of transport, Repair of software, Maintenance of medical equipment, Personal Computer and office equipment maintenance, telephone maintenance, Vehicle maintenance and servicing, Software maintenance, Fire extinguisher maintenance, Transport rental, Medical equipment rental. |
| Medical Equipment, Services, and Related Costs | Expenses for business trips, Training and advanced education (excluding drivers), Driver training, External medical care services (excluding diagnostic tests and investigations), Communications and internet, Telemedicine services, Laundry services, Catering services, Insurance (excluding transport and premises), Motor vehicle insurance, Car maintenance and servicing, Cleaning services (excluding premises), Accounting and bookkeeping, Auditing, Occupational safety, Pest control and disinfection, Waste management, Disability assessment and certification, Processing of medical records and document archiving (annually), Bank loan services, Environmental protection services, Cryptographic protection of information license, Banking services, Reimbursement for preferential medicines and insulin, Production of electronic keys, Information and consulting services, Documentation preparation, Legal services, Refuelling card readers, Verification of medical devices, Verification of scales, Maintenance services for cash registers, State registration of information/changes in the Unified State Register of Enterprises and Organizations of Ukraine, Providing access to the online service "Record Portal" “RIMS” "Medstar", Radio broadcasts, Standard service package for "Medoc" software, Support for the "TIS-Salary" program, Data processing and generation of a qualified public key certificate, Support for the "Accounting of Medical Personnel" software package, Support for the "Medical Statistics" software program, Height meter, Medical measuring tape, Disposable consumables for electrocardiograph, Electrocardiogram paper, Consumables for peak flow meter, Neurological hammer, Glucometer, Test strips, Disposable lancets, Spatulas, Paper towels for medical purposes, Napkins (wet/dry), Disposable couch sheets, Syringes and needles, Disposable infusion systems, Venous catheters, Other catheters, Vacuum tubes (vacutainers), Dressings (including tampons, cotton wool, bandages), Disposable small surgical sets, Disposable examination instruments, Laboratory glassware, Dispensers, Atraumatic needles with suture thread, Needle holder, Haemostatic clamp, Double-sided button probe, Korentang, Vertically curved surgical scissors, Straight surgical scissors, Anatomical tweezers, Surgical tweezers, Disposable scalpels or blades with handle, Steam steriliser, Retaining bandages, Support and compression dressings, Plasters/adhesive tapes, Surgical absorbents, Reagents/chemicals, Medical paper, Non-vacuum test tubes, Urine containers, Thermometers (including digital and infrared), Bactericidal lamp, Tripod/holder, Pipettes, Nappies, Faecal colostomy bags, Urological pads, Consumables for COVID-19 biological material collection, Consumables for biological specimen collection, Haematology analyser, Biochemistry analyser, Electrocardiograph, Centrifuge, Examination couch (including gynaecological chair), Oxygen concentrator, Stethoscope, Medical screen, Paediatric scales, Adult scales, Stethophonendoscope, Blood pressure monitor with various cuffs, Portable pulse oximeter, Ophthalmoscope, Medical torch, Peak flow meter, Spirometer, Visual acuity testing tables, Blood glucose testing kit (glucometer, strips, lancets, gloves), Instrument containers, Consumable containers, General-purpose containers and trays, Nebuliser, Manual ventilation bag (ambu type), Medical stretchers, Wheelchair, Portable shadowless illuminator, Reusable small surgical sets with steriliser, Reusable examination instruments (steriliser included), Doctor's/nurse's bag, Cool bag with cold storage batteries, Medicine refrigerator, Vaccine refrigerator, Microscope, Lancet, Tourniquet, X-ray viewer, Defibrillator, Ear syringe equipment, Urine analyser, Mobile instrument table, Mobile changing table, Changing table (for waiting room), Medicine and medical device storage cabinet, Labelled containers for healthcare waste separation, Surgical scissors, Dental chair, Bed, Medical gowns (workwear), Oxygenator for adults over 50 kg with a set of main pipes, Mechanical heart valve, Sterility indicator "Sterilan" 180/60, Medical gauze, Sterile operating cover (120/80). |
| Medical equipment depreciation | Height meter, Medical measuring tape, Disposable consumables for electrocardiograph, Electrocardiogram paper, Disposable consumables for peak flow meter, Neurological hammer, Glucometer, Test strips, Disposable lancets, Spatulas, Medical paper towels, Napkins (including wet), Disposable couch sheets, Syringes and needles, Disposable infusion systems, Venous catheters, Other catheters, Vacuum tubes (vacutainers), Dressings (including tampons, cotton wool, bandages), Disposable small surgical sets, Disposable examination instruments, Laboratory glassware, Dispensers, Atraumatic needles with suture thread, Needle holder, Haemostatic clamp, Double-sided button probe, Korentang, Vertically curved surgical scissors, Straight surgical scissors, Anatomical tweezers, Surgical tweezers, Disposable scalpels or scalpel blades with handle, Steam steriliser, Retaining bandages, Support and compression dressings, Plasters/adhesive tapes, Surgical absorbents, Reagents/chemicals, Medical paper, Non-vacuum test tubes, Urine containers, Thermometers (including digital and infrared), Bactericidal lamp, Tripod/holder, Pipettes, Nappies, Faecal colostomy bags, Urological pads, Consumables for COVID-19 biological material collection, Consumables for biological specimen collection, Haematology analyser, Biochemistry analyser, Electrocardiograph, Centrifuge, Examination couch (including gynaecological chair), Oxygen concentrator, Stethoscope, Medical screen, Children's scales, Adult scales, Stethophonendoscope, Blood pressure monitor with various cuffs, Portable pulse oximeter, Ophthalmoscope, Medical torch, Peak flow meter, Spirometer, Visual acuity testing tables, Blood glucose testing kit (glucometer, strips, lancets, gloves), Instrument containers, Consumable containers, General-purpose containers and trays, Nebuliser, Manual ventilation bag (ambu type), Medical stretchers, Wheelchair, Portable shadowless illuminator, Reusable small surgical sets with steriliser, Reusable examination instruments (steriliser provided), Doctor's/nurse's bag, Cool bag with cold storage batteries, Medicine refrigerator, Vaccine refrigerator, Microscope, Lancet, Tourniquet, X-ray viewer, Defibrillator, Ear syringe equipment, Urine analyser, Mobile instrument table, Mobile changing table, Changing table (for waiting room), Medicine and medical device storage cabinet, Labelled containers for healthcare waste separation, Surgical scissors, Dental chair, Bed, Medical gowns (workwear), Bactericidal irradiator, Gynaecological chair, Oxygenator for adults over 50 kg with main pipes, Mechanical heart valve, Sterility indicator, Medical oilcloth, Sterile operating cover, Needle disposer, Alcohol analyser, Hygrometer, Oxygen pillow, Sterilisation box. |
| Costs Associated with the Operation of Transportation Facilities | All-wheel drive passenger car, Passenger car, Motorcycle, All Terrain Vehicle, Scooter, Bicycle, First aid kits, Spare parts for transportation vehicles, Spare parts for transportation vehicles, Automotive tools (screwdriver, wrench, hammer, etc.), Fuels and lubricants for transportation (including gas). |
| Costs associated with the operation of computer equipment | Personal computer (all-in-one, laptop, tablet, etc.), All-in-one printer (or printer + scanner), Individual Personal Computer parts (monitor, system unit), Modem/Router/Wi-Fi unit, Universal Serial Bus (USB) flash drives, Batteries, Calculator, Printer cartridges, Appliance care products (cleaning wipes/sprays, etc.), Toner, Mouse, Keyboard. |
| Costs associated with the operation of other non-medical equipment | Telephone (Internet Protocol, mobile, landline and other means of communication), Switch, Air conditioner, Generator, Water heater/Boiler, Convector, Microwave oven, Television, Pump, Stove (gas, electric), Iron, Refrigerator, Hand tools, Power tools (chainsaw, hammer drill, drill, grinder, etc.), Heater (electric fireplace, other), Washing machine. |
| Depreciation of Transportation Vehicles | Passenger car (all-wheel drive), Passenger car, Motorcycle, Quad bike, Scooter, Bicycle, First aid kits, Spare parts for transportation vehicles, Automotive tools (screwdriver, wrench, hammer, etc.). |
| Software depreciation | Software, Application software (except for primary care), Specialised application software for primary care. |
| Depreciation of computer equipment | Personal computer (all-in-one, laptop, tablet, etc.), All-in-one (or printer + scanner), Individual parts of the Personal Computer (monitor, system unit), Modem/router/Wi-Fi unit, Universal Serial Bus (USB) flash drives, Batteries, Calculator, Printer Cartridge, Appliance care products (cleaning wipes/sprays, etc.), Toner, Mouse, Keyboard. |
| Depreciation of other non-medical equipment | Telephone (Internet Protocol, mobile, landline and other means of communication), Switch, Air conditioner, Generator, Water heater/Boiler, Convector, Microwave oven, Television, Pump, Stove (gas, electric), Iron, Refrigerator, Hand tools, Power tools (chainsaw, hammer drill, drill, grinder, etc.), Heater (electric fireplace, other), Washing machine, Server cabinet, Webcam, Speakers, Uninterruptible power supply, Electric kettle. |
| Depreciation of household equipment | Metal bicycle parking, Mosquito net, Extension cords, Video recorder, Fire extinguishers, Set of car covers, white boards. |
| Furniture depreciation | Staff desks, Office chairs and armchairs, Benches and corridor banquettes, Filing and clothing cabinets, Workwear storage cabinets, Safes, Other furniture (kitchen, waiting area, seating area), Bedside tables. |

Table S4: Median and inter-quartile (IQR) of cost-per-registered person of Primary Healthcare providers across the explanatory variables of the study.

|  |  | *Cost-per-registered person* | |
| --- | --- | --- | --- |
| Variable | *n* | *Median*  *(USD)* | *Interquartile Range*  *(USD)* |
| **Locality**  Urban  Rural | 81  19 | 49.81  55.24 | 34.37, 52.74  44.36, 65.01 |
| **Type of ownership**  Public  Private | 70  30 | 52.28 19.77 | 49.80, 56.29  12.93, 30.87 |
| **Region**  North  South  East  West  Central | 9  17  7  35  32 | 58.44 17.82 51.19 50.09 50.83 | 51.73, 68.98  12.83, 45.36  13.37, 53.78  43.92, 55.24  44.22, 54.27 |
| **Oblast**  Cherkaska  Chernihivska  Chernivetska  Dnipropetrovska  Donetska  Ivano-Frankivska  Kharkivska  Khmelnytska  Kirovohradska  Kyiv city  Kyivska  Lvivska  Mykolaivska  Odeska  Poltavska  Rivnenska  Sumska  Ternopilska  Vinnytska  Volynska  Zhytomyrskа | 3  1  6  3  2  1  1  6  2  7  4  4  2  15  2  10  5  3  12  5  6 | 49.80 51.73 47.88 52.56 32.62 65.01 26.58 50.02 38.68 51.00 36.59 48.81 29.15 17.82 57.63 52.50 69.00 40.22 50.64 50.36 53.39 | 46.13, 55.50  51.73, 51.73  43.92, 50.38  13.37, 54.67  11.47, 53.78  65.01, 65.01  26.58, 26.58  46.88, 74.50  26.16, 51.19  44.08, 53.16  19.21, 57.20  43.84, 59.59  12.93, 45.36  12.75, 46.65  56.82, 58.44  34.37, 55.28  67.47, 79.00  30.87, 43.39  43.40, 53.08  50.09, 54.69  50.31, 57.87 |
| **Mountain region**  Non-mountain  Mountainous | 97  3 | 50.29  64.55 | 34.37, 54.24  40.50, 65.01 |
| **Service location points**  1  2-10  11-20  >20 | 31  29  19  21 | 20.64  50.65  54.69  50.64 | 12.93, 36.65  47.63, 55.24  51.19, 60.02  50.09, 54.30 |
| **Area of each PHC provider (m^2^)**  0-2,000  2,001-4,000  >4,000 | 44  16  40 | 30.77  52.57  51.46 | 16.35, 47.33  50.20, 57.08  49.94, 54.98 |
| **Average monthly number of registered persons**  0-10,000  10,001-20,000  20,001-30,000  30,001-40,000  >40,000 | 44  11  12  13  20 | 30.77  53.78  52.21  50.38  50.64 | 16.35, 53.16  48.13, 57.87  50.36, 57.34  46.88, 54.69  49.63, 52.66 |
| **Average number of doctors with registered persons**  0-10  11-20  21-30  >31 | 46  21  15  18 | 31.92  52.15  50.58  50.83 | 17.78, 53.78  50.09, 56.82  49.81, 54.69  49.46, 52.74 |
| **Average number of registered persons per doctor**  <1800 registered persons (optimal)  >1800 (greater than optimal) | 97  3 | 50.09  55.50 | 34.37, 54.24  52.40, 76.21 |
| **Average number of employees**  0-30  31-60  61-90  >90 | 35  13  12  40 | 21.62  57.87  50.89  51.09 | 13.30, 40.50  52.40, 67.47  46.92, 53.16  49.95, 54.68 |
| Abbreviations: PHC: Primary Healthcare | | | |

Table S5: Univariate regression model summary for the relationship between the explanatory variables and cost-per-registered person.

| Variable | *ᵝ* | *95% Confidence*  *Interval* | *Standard error* | *t-statistic* | *p-value* | *R-squared* | *Adjusted*  *R-squared* | *F-statistic (d.f.)* |
| --- | --- | --- | --- | --- | --- | --- | --- | --- |
| **Locality**  Urban  Rural | 0 (base) 11.27 | 2.16, 20.38 | 4.59 | 2.46 | 0.016 | 0.06 | 0.05 | F(1,98) = 6.03 |
| **Type of ownership**  Public  Private | 0 (base)  -31.47 | -36.44, -26.50 | 2.51 | -12.56 | <0.001 | 0.62 | 0.61 | F(1,98) = 157.71 |
| **Region**  North  South  East  West  Central | 0 (base)  -34.90  -24.27  -12.29  -14.01 | -47.98, -21.82  -40.27, -8.28  -24.15, -0.43  -25.98, -2.03 | 6.59  8.06  5.97  6.03 | -5.30  -3.01  -2.06  -2.32 | <0.001  0.003  0.042  0.022 | 0.28  0.28  0.28  0.28 | 0.25  0.25  0.25  0.25 | F(4,95) = 9.24  F(4,95) = 9.24  F(4,95) = 9.24  F(4,95) = 9.24 |
| **Mountain region**  Non-mountain  Mountainous | 0 (base) 11.58 | -9.88, 33.03 | 10.81 | 1.07 | 0.29 | 0.01 | 0.002 | F(1,98) = 1.15 |
| **Number of branches per PHC provider** | 0.58 | 0.33, 0.83 | 0.13 | 4.57 | <0.001 | 0.18 | 0.17 | F(1,98) = 20.89 |
| **Average number of monthly registered persons** | 0.00014 | 0.000025, 0.00026 | 0.000058 | 2.41 | 0.018 | 0.06 | 0.05 | F(1,98) = 5.79 |
| **Average number of doctors with registered persons** | 0.20 | 0.04, 0.35 | 0.08 | 2.51 | 0.014 | 0.06 | 0.05 | F(1,98) = 6.29 |
| **Average number of registered persons per doctor** | 0.01 | 0.00, 0.02 | 0.005 | 2.56 | 0.012 | 0.06 | 0.05 | F(1,98) = 6.54 |
| **Average number of employees** | 0.10 | 0.05, 0.14 | 0.02 | 4.15 | <0.001 | 0.15 | 0.14 | F(1,98) = 17.25 |
| **Area of PHC Provider** | 0.0015 | 0.00076, 0.0021 | 0.00035 | 4.16 | <0.001 | 0.15 | 0.14 | F(1,98) = 17.29 |
| Abbreviations: d.f.: Degrees of Freedom; PHC: Primary Healthcare | | | | | | | | |

Table S6: Generalized-linear model summary (with a gamma distribution) of the final model specification for cost-per-registered person after adjustment for confounding explanatory variables.

| Variable | *Coefficient** | *95% Confidence*  *Interval* | *Standard*  *error* | *z-statistic* | *p-*  *value* |
| --- | --- | --- | --- | --- | --- |
| **Locality**  Urban  Rural | 0 (base)  0.12 | -0.03, 0.27 | 0.08 | 1.56 | 0.118 |
| **Type of ownership**  Public  Private | 0 (base)  -0.98 | -1.15, -0.82 | 0.08 | -11.71 | <0.001 |
| **Average number of registered persons per doctor** | -0.00025 | -0.00045, -0.000055 | 0.0001 | -2.51 | 0.012 |
| **Mountain region**  Non-mountain  Mountainous | 0 (base)  0.54 | 0.20, 0.88 | 0.17 | 3.15 | 0.002 |
| **Average number of doctors with registered persons** | -0.00225 | -0.0051, 0.00057 | 0.0014 | -1.57 | 0.117 |
| **Region**  North  South  East  West  Central | 0 (base)  -0.39  -0.23  -0.17  -0.13 | -0.62, -0.15  -0.52, 0.05  -0.37, 0.04  -0.34, 0.08 | 0.12  0.15  0.10  0.11 | -3.22  -1.61  -1.60  -1.23 | 0.001  0.108  0.110  0.217 |
| **Residual d.f. = 90**  **Scale parameter = 0.07**  **(1/d.f.) Deviance = 0.08**  **(1/d.f.) Pearson = 0.07**  **Log likelihood = -473.07**  **AIC = 966.15**  **BIC = 992.20** | |  |  |  |  |
| * Adjusted for all other variables in the table  Abbreviations: AIC: Akaike Information Criterion; BIC: Bayes Information Criterion; d.f.: Degrees of Freedom; | | | | | |

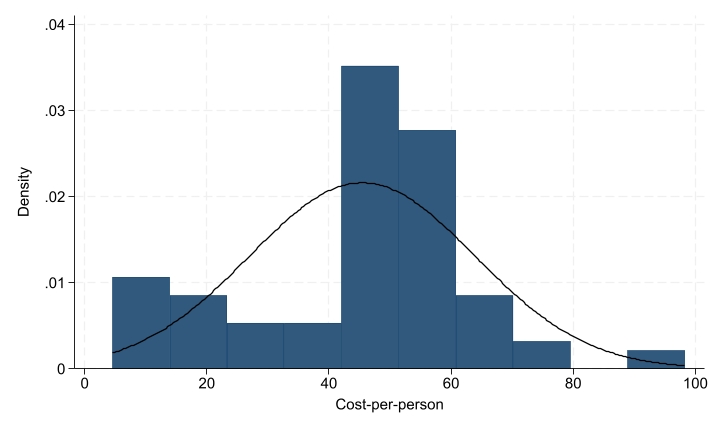

### Supplemental Figures

Figure S1: Frequency distribution of cost-per-person superimposed with a normal distribution.

This graph compares the frequency distribution of the cost-per-registered person data, in the form of a histogram, to that of a theoretical normal distribution. The deviations from the normal distribution curve indicate that the cost-per-registered person data does not follow a normal distribution.

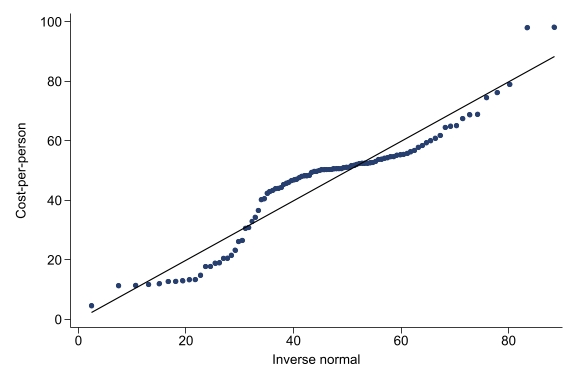

Figure S2: Q-Q plot of cost-per-person showcasing deviations from normal distribution.

This graph compares the quantiles of the cost-per-registered person data to those of a theoretical normal distribution in the form of a Q-Q plot. The deviations from the diagonal normal distribution line indicate that the cost-per-registered person data does not follow a normal distribution.
